# Complement activation induces excessive T cell cytotoxicity in severe COVID-19

**DOI:** 10.1101/2021.06.08.21258481

**Authors:** Philipp Georg, Rosario Astaburuaga-García, Lorenzo Bonaguro, Sophia Brumhard, Laura Michalick, Lena J. Lippert, Tomislav Kostevc, Christiane Gäbel, Maria Schneider, Mathias Streitz, Vadim Demichev, Ioanna Gemünd, Matthias Barone, Pinkus Tober-Lau, Elisa Theresa Helbig, Julia Stein, Hannah-Philine Dey, Daniela Paclik, Michael Mülleder, Simran Kaur Aulakh, Henrik E. Mei, Axel R. Schulz, Stefan Hippenstiel, Victor Max Corman, Dieter Beule, Emanuel Wyler, Markus Landthaler, Benedikt Obermayer-Wasserscheid, Peter Boor, Münevver Demir, Hans Wesselmann, Norbert Suttorp, Alexander Uhrig, Holger Müller-Redetzky, Jacob Nattermann, Wolfgang M. Kuebler, Christian Meisel, Markus Ralser, Joachim L. Schultze, Anna C. Aschenbrenner, Charlotte Thibeault, Florian Kurth, Leif-Erik Sander, Nils Blüthgen, Birgit Sawitzki

## Abstract

Severe COVID-19 is linked to both dysfunctional immune response and unrestrained immunopathogenesis, and it remains unclear if T cells also contribute to disease pathology. Here, we combined single-cell transcriptomics and proteomics with mechanistic studies to assess pathogenic T cell functions and inducing signals. We identified highly activated, CD16^+^ T cells with increased cytotoxic functions in severe COVID-19. CD16 expression enabled immune complex-mediated, T cell receptor-independent degranulation and cytotoxicity not found in other diseases. CD16^+^ T cells from COVID-19 patients promoted microvascular endothelial cell injury and release of neutrophil and monocyte chemoattractants. CD16^+^ T cell clones persisted beyond acute disease maintaining their cytotoxic phenotype. Age-dependent generation of C3a in severe COVID-19 induced activated CD16^+^ cytotoxic T cells. The proportion of activated CD16^+^ T cells and plasma levels of complement proteins upstream of C3a correlated with clinical outcome of COVID-19, supporting a pathological role of exacerbated cytotoxicity and complement activation in COVID-19.

## Introduction

Severe acute respiratory distress syndrome Coronavirus 2 (SARS-CoV-2) infection in humans causes a diverse spectrum of clinical manifestations, ranging from asymptomatic disease to acute respiratory distress syndrome (ARDS) and multi-organ failure (Miyazawa, 2020).

In addition to direct virus-induced injury to the respiratory system and other organs, increasing evidence suggests that the immune response evoked by SARS-CoV-2 infection contributes to the pathophysiology of Coronavirus disease (COVID-19), particularly during severe disease courses (Gustine and Jones, 2021; McKechnie and Blish, 2020; Vabret et al., 2020).

Both, conventional CD4^+^ T helper cells and particularly CD8^+^ Cytotoxic T Lymphocytes (CTL), play important roles in the control of respiratory viral infections. Not surprisingly, T cell responses have been discussed to play a protective role in COVID-19 (Jacob, 2020). High numbers of SARS-CoV-2-specific CD4^+^ and CD8^+^ T cells have been associated with a milder disease course. While this had been interpreted as a predominantly protective role of T cell responses during COVID-19 (Rydyznski Moderbacher et al., 2020), complementary data do not unequivocally support this idea (Feng et al., 2020; Mathew et al., 2020; Peng et al., 2020; Thieme et al., 2020). The extent of SARS-CoV-2-specific T cell responses could not be directly linked to disease severity, with high T cell numbers not necessarily translating into mild courses of COVID-19. In fact, the number of SARS-CoV-2-specific CD4^+^ and CD8^+^ T cells were found to be comparable or even higher in COVID-19 patients displaying severe versus mild courses of disease (Feng et al., 2020; Mathew et al., 2020; Peng et al., 2020; Thieme et al., 2020).

Along this line, flow cytometric analysis of 19 COVID-19 patients revealed a higher state of T cell activation in all T cell compartments (CD4^+^, CD8^+^, Double negative (DN)) (Zenarruzabeitia et al., 2021).

Interstitial T cell infiltration is commonly observed in pathological specimens of COVID-19 pneumonia along with macrophage accumulation in the alveolar space, and it has been hypothesized that T cell infiltrates also contribute to damaging alveolar walls and endothelial cell injury known as lymphocytic endotheliitis (Miyazawa, 2020; Varga et al., 2020).

All this argues for a complex relationship between T cell immune responses and disease outcome during COVID-19 beyond a mere quantitative influence of the T cell compartment with additional factors shaping the quality of T cell responses and consequently ensuing pathology as well as disease amelioration. For a better understanding of the disease it is therefore important to identify whether and which T cells have a pathogenic role. In addition, the mechanism by which they are induced and act need to be revealed as studies in this direction are currently lacking (Yan et al., 2021).

Here, we combined single-cell proteomics and transcriptomics with mechanistic studies to reveal alterations in the T cell compartment and their causative signals and functional relevance, which explains important immunopathological features observed in severe COVID-19. Mass cytometry (CyTOF) and single-cell RNA-seq combined with VDJ-seq-based T cell clonotype identification were used to determine COVID-19-as well as severity-specific alterations in the T cell compartment within two cohorts. In addition to the severity-independent formation of highly activated HLA-DR^hi^CD38^hi^CD137^+^Ki67^+^ T Follicular Helper (TFH) -like cells and CD8^+^ cytotoxic T lymphocytes (CTLs) in COVID-19, we describe a C3a-driven specific induction of activated CD16 expressing cells in severe COVID-19 patients in the three main T cell compartments. These T cells display increased immune complex-mediated, TCR-independent cytotoxicity causing activation and release of chemokines by lung endothelial cells. This mechanism most likely contributes to the profound organ pathophysiology and endotheliitis observed in autopsy samples of severe COVID-19 patients.

## Results

### Profound T cell activation and induction of CD16 expressing CD4^+^, CD8^+^ TCRab^+^ and TCRgd^+^ T cells in severe COVID-19

To reveal putative immunopathogenic roles of T cells in severe COVID-19 we performed mass cytometry of whole blood samples from mild and severe COVID-19 patients during the acute and convalescent phase, patients suffering from other acute respiratory infections (Flu-like illness, FLI), patients chronically infected by human immunodeficiency virus (HIV) or hepatitis B (HBV) as well as controls (Figure 1A). We reported previously that T cell numbers were reduced in COVID-19 patients during acute infection compared to controls (Schulte-Schrepping et al., 2020). To further interrogate the T cell space, acquired T cells (CD45^+^CD3^+^CD19^-^CD15^-^) were pre-gated into CD4^+^ T helper cells (CD3^+^, CD8^-^TCRgd^-^), CD8^+^ CTLs (CD3^+^, CD8^+^TCRgd^-^), and TCRgd^+^ (CD3^-^, CD8^-^ TCRgd^+^) cells. As expected, samples from HIV patients showed an inverted CD4/CD8 ratio, whereas the proportions of all three main T cell compartments were similar between all other patient groups (mild and severe COVID-19, FLI, HBV) and controls (median levels: T helper cells 63.3-71.5%, CTLs 27.2-34.8%, TCRgd^+^ 0.9-2.9%). Unsupervised clustering analysis using 29 surface antigens and the proliferation marker Ki67 partitioned the pre-gated T helper cells, CTLs, and TCRgd^+^ T cells into 18, 15 and 14 individual cell clusters, respectively (Figure 1B-D). A significant proportion of T cells from acutely infected mild and severe COVID-19 patients were clearly separated in UMAP space from those of the other patient groups or controls (Figure 1B).

**Figure 1.**
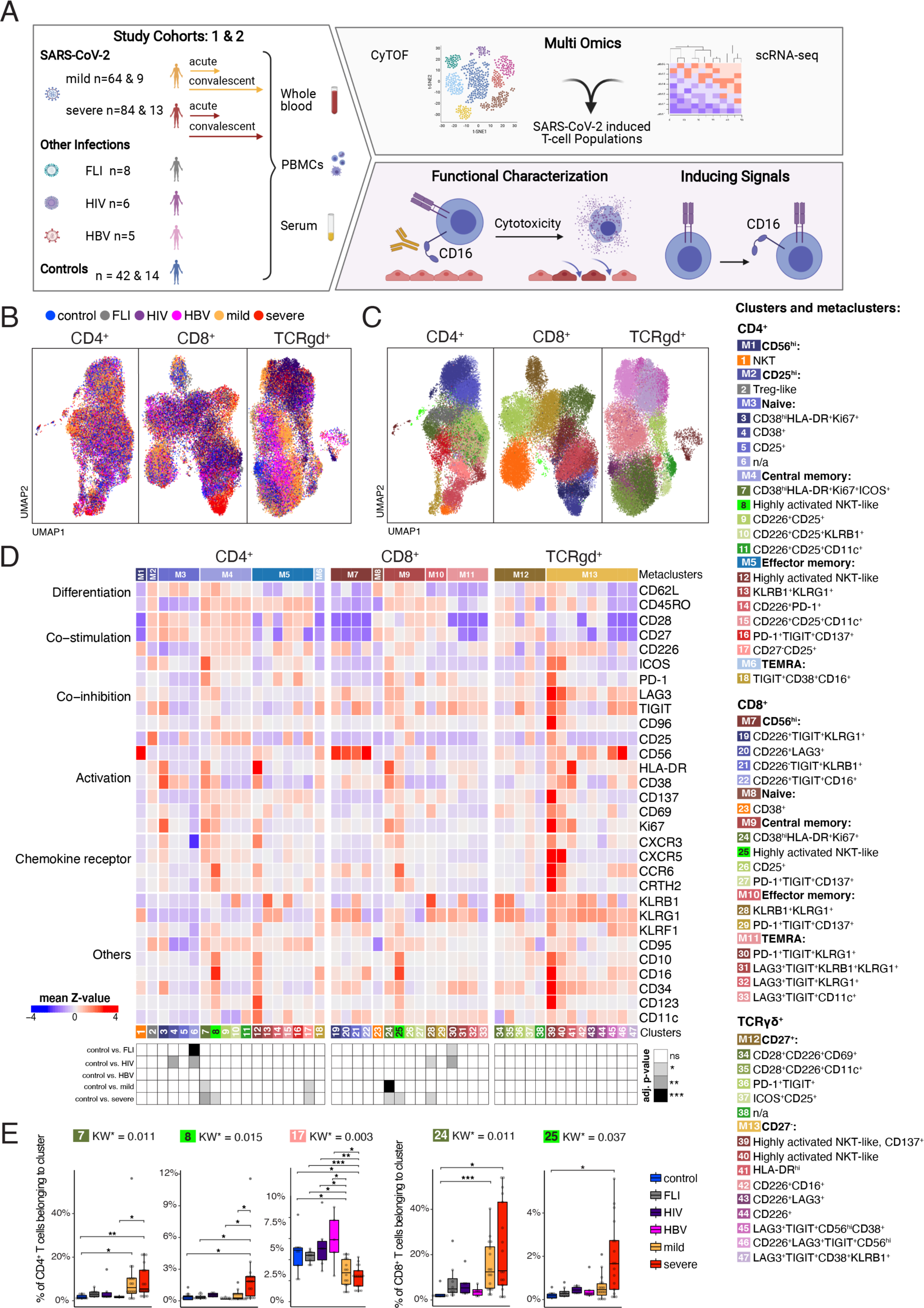
Accumulation of HLA-DR^hi^CD38^hi^ highly activated but also CD16 expressing CD4^+^ and CD8^+^ T cells in severe COVID-19. **A,** Overview of the study cohort and methodological pipeline. Blood samples were collected from mild and severe COVID-19 patients during the acute and convalescent phase enrolled in Berlin (cohort 1) or Bonn (cohort 2), patients suffering from other acute respiratory infections (FLI), being chronically infected by human immunodeficiency virus (HIV) or hepatitis B (HBV) as well as controls. Mass cytometry (CyTOF) and single-cell RNA-seq combined with VDJ-seq-based T cell clonotype identification were used to determine COVID-19 as well as severity-specific alterations in the T cell compartment. The obtained results in combination with additional serum proteomics data and clinical information were used to develop hypotheses on their functional properties and mechanisms leading to their differentiation, which were tested in *ex vivo* cultures. Detailed sample information included in all reported assays can be found in **Table S1**. **B,** UMAPs generated of CD4^+^ (left), CD8^+^ (middle), and TCRgd^+^ (right) T cells from mass cytometry data. Cells are colored according to donor origin (blue, age-matched controls; gray, FLI; violet, HIV; pink, HBV; yellow, mild COVID-19 acute phase; red, severe COVID-19 acute phase). For visualization purposes, each UMAP shows 28000 cells. **C,** UMAPs generated of CD4^+^ (left), CD8^+^ (middle), and TCRgd^+^ (right) T cells from CyTOF data. Cells are colored according to the cell cluster origin. For visualization purposes, each UMAP shows 28000 cells. **D,** Heatmap of CyTOF data (described antibody panel, cohort 1) covering CD4^+^ (left panel), CD8^+^ (middle panel) and TCRgd^+^ (right panel) T cells. Z-score standardized staining intensity of each marker (rows) per cluster (1 to 47, in columns, lower part). Clusters were grouped into metaclusters, as defined by the numbers 1 to 13 (in columns, upper part). Significance levels of differential cluster frequency for the following groups: controls (n = 9), FLI (n = 8), HIV (n = 6), HBV (n = 5), mild acute COVID-19 (n = 24) and severe acute COVID-19 (n = 29). All COVID-19 and FLI samples were collected between days 4 and 13 post-symptom onset ( = first day of sample collection per patient). The cluster abundance was tested via adjusted Dunn’s post-hoc test (Benjamini-Hochberg) for clusters with significant adjusted (Benjamini-Hochberg) Kruskal-Wallis test (adjusted p-value < 0.05). All combinations where tested, only comparisons with healthy controls are shown. **E,** Box plots of CD4^+^ (7, 8, 17) and CD8^+^ (24, 25) T cell clusters determined by mass cytometry as defined in the legend (whole blood, cohort 1) generated from controls (n = 9), FLI (n = 8), HIV (n = 6), HBV (n = 5), mild acute COVID-19 (n = 16) and severe acute COVID-19 (n = 17) patient samples. KW* shows the adjusted p-value (Benjamini-Hochberg) of a Kruskal-Wallis test. The abundance of each cluster was compared between severity groups via adjusted Dunn’s post-hoc test (Benjamini-Hochberg) for clusters with KW* < 0.05. All combinations where tested, only comparisons with healthy controls are shown. (*p<0.05, **p < 0.01,***p < 0.001).

Samples of COVID-19 patients collected during the first three weeks after symptom onset independent of the severity compared to samples from patients with other infections or controls were characterized by increased proportions of CD4^+^ T helper cell cluster 7 (CD38^hi^HLA-DR^+^Ki67^+^ICOS^+^), whereas the abundance of cluster 17 (CD27^-^CD25^+^) was lower (Figure 1D, E). Cluster 7 T cells are characterized by high expression of activation markers such as HLA-DR, CD38, CD137, CD69, and the proliferation marker Ki67. Furthermore, the T cells in this cluster do express CXCR5, ICOS, and PD-1, resembling TFH-like cells. This increase was not seen for patients with other acute respiratory infections (FLI) or chronic viral infections (HIV or HBV). Interestingly, T helper cells in severe COVID-19 patients showed higher proportions of cluster 8 T cells (CD4^+^, highly activated Natural Killer T cell (NKT)-like), which in addition to the expression of CD38, HLA-DR, CD137, CD69, Ki67 and CXCR3, were characterized by high levels of CCR6 and CD16. Those T cells belong to the CD62L^+^CD45RO^+^ central memory metacluster, reflecting a more recent differentiation. In contrast, in patients suffering from chronic HIV or HBV infection we observed a tendency towards a higher proportion of cluster 18 (TIGIT^+^CD38^+^CD16^+^). T cells from this cluster were classified as terminally differentiated RA^+^ cells, expressed low levels of CD16, and showed no signs of recent activation such as CD137 expression

Surprisingly, this pattern of activation states was not restricted to the CD4^+^ T cell compartment, but also detectable in CD8^+^ CTLs and TCRgd^+^ T cells. Both mild and severe COVID-19 patients displayed increased proportions of cluster 24 T cells (CD8^+^CD38^hi^HLA-DR^+^Ki67^+^) in comparison to the other patient groups (Figure 1D, E). Severe COVID-19 was further characterized by increased abundance of cluster 25 T cells (CD8^+^, highly activated NKT-like), also expressing high levels of CCR6 and CD16. The existence of a CD3^+^CD16^+^HLA-DR^+^ T cell population was confirmed by flow cytometry in a second cohort of COVID-19 patients (Figure S1A). In contrast, cluster 30 T cells (PD-1^+^TIGIT^+^KLRG1^+^) co-expressing CD137 and several co-inhibitory receptors (PD-1, TIGIT, LAG3) were elevated in samples from patients with other acute respiratory infections (FLI) or HIV infection. In chronic HIV and HBV infection, we observed a tendency towards a higher proportion of cluster 32 cells (LAG3^+^TIGIT^+^KLRG1^+^) containing terminally differentiated CD16 expressing T cells (data not shown).

Time-dependent analysis of the CD4^+^ and CD8^+^ T cell clusters revealed a trend to faster accumulation of the activated CD38^hi^HLA-DR^+^Ki67^+^ expressing clusters 7 and 24 during the first week after SARS-CoV-2 infection in severe versus mild COVID-19 (Figure S1B, C). In contrast, the highly activated CD16 expressing NKT-like CD4^+^ and CD8^+^ T clusters appeared only during the second week of infection.

Collectively, our overall findings concerning the identification of circulating highly activated and proliferating T helper cells and CTLs after SARS-CoV-2 infection is in line with recent reports (Mathew et al., 2020; Rydyznski Moderbacher et al., 2020; Stephenson et al., 2021). However, and strikingly, we uncovered a unique activation and differentiation program in a subset of T cells across all three major T cell compartments (T helper cells, CTL and TCRgd^+^ T cells), characterized by CD16 and CCR6 expression in severe COVID-19.

### Single-cell transcriptomics reveal shift towards disproportionally high cytotoxic and degranulation potential of T cells in severe COVID-19

To reveal functional information on the COVID-19- and severity-specific T cell clusters, we performed single-cell RNA-seq analysis of peripheral blood mononuclear cell (PBMC) samples as well as purified CD38 expressing T cells from acutely infected and convalescent mild and severe COVID-19, as well as FLI, HBV and controls. Cluster analysis of the whole T cell space from PBMC and CD38^+^ T cell libraries generated 15 different T cell clusters and one unspecified cell cluster (Figure 2A, B). To align the CyTOF and scRNA-seq T cell clusters, we applied a feature-based cluster annotation approach. The selected feature list contained genes for all markers analysed in CyTOF measurements plus additional genes such as T cell state-specific transcription markers (Figure 2B). T cell clusters were composed of solely *CD4* expressing T cell clusters 1 to 7, mixed *CD4*, *CD8A*/*B* & *TRGC2* expressing T cell clusters 8 and 9, mixed *CD8A*/*B* & *TRGC2* expressing T cell clusters 10 to 13 as well as solely *CD8* transcribing T cell clusters 14 & 15. Cluster 7, 8 and 10 were characterized by highly activated T cells with high transcription of *CD38* and *HLA-DR* genes (Figure 2B, C). Cluster 7 T cells showed the typical features of activated TFH cells, as in addition to *CD38* and *HLA-DR* genes, they transcribed *ICOS*, *CD40LG*, *PDCD1* (PD-1) and *CXCR5* (Figure 2B). Cluster 8 T cells transcribed the highest *MKI67* (Ki67) levels, indicative of the highest proliferative potential (Figure 2B, C). In addition, they expressed *FCGR3A*, the gene encoding for CD16A, which was even more pronounced for cluster 10 T cells (Figure 2B, C). The proportion of T cells belonging to cluster 7, 8 and 10 was higher in COVID-19 patients compared to FLI or HBV patients as well as controls (Figure 2D, E). We observed other T cell clusters with *FCGR3A* expression (cluster 9, 11, 12, 13). Only cluster 9 T cells transcribed *SELL* (CD62L) and displayed a central memory phenotype, whereas cluster 11, 12 and 13 T cells displayed a more advanced differentiation profile. Overall, cluster 7 contains TFH-like cells similar to the CyTOF cluster 7, whereas cluster 8, 9 and 10 most likely contain a mixture of highly activated and CD16^+^ NKT-like cells with similarities to the CyTOF clusters 8, 24 and 25. Next, we performed a Gene Ontology (GO) enrichment analysis across clusters 7, 8, 9 and 10, comparing mild and severe COVID-19 T cells (Figure 2F). Interestingly, whereas T cells from mild COVID-19 patients showed an enrichment for cellular responses to type I interferon and antiviral defense, we observed a very specific enrichment of genes involved in degranulation in severe COVID-19 T cells. This selective enrichment was validated by a Gene Set Enrichment Analysis (GSEA) for genes belonging to the GO terms ‘response to type I interferon’ and ‘defense response to virus’, which were enriched in mild, and for genes belonging to a cytotoxicity signature, which were enriched in severe COVID-19 T cells (Figure 2G). Indeed, several genes known to promote degranulation (*LAMP1*, *STX11*) or exerting cytotoxic potential (*PRF1*, *GZMB*, *GZMH*, *GZMK*) displayed a tendency towards higher expression in severe COVID-19 across clusters 7, 8, 9 & 10 T cells (Figure 2H+S2B) or the whole T cell space (Figure S2C). Importantly, we validated these findings in scRNA-seq data from our second cohort (Figure S3). Applying the same feature selection-based annotation approach to the second cohort resulted in identification of COVID-19-specific T cell clusters, which consistently aligned to the ones from the first cohort (Figure S3A-D). Cluster 6 and 7 contained CD4^+^ T cells transcribing *ICOS*, *CD40LG*, *PDCD1* and *CXCR5* and thus resembled cluster 7 T cells from cohort 1, although the expression level of *CD38* and *HLA-DR* genes seemed to be lower. Clusters 10 & 13 from the second cohort resembled cluster 10 & 8 in cohort 1, respectively. Furthermore, GSEA similarly showed increased expression of genes mediating cytotoxicity in severe COVID-19 T cells across these clusters (Figure S3E, F).

**Figure 2.**
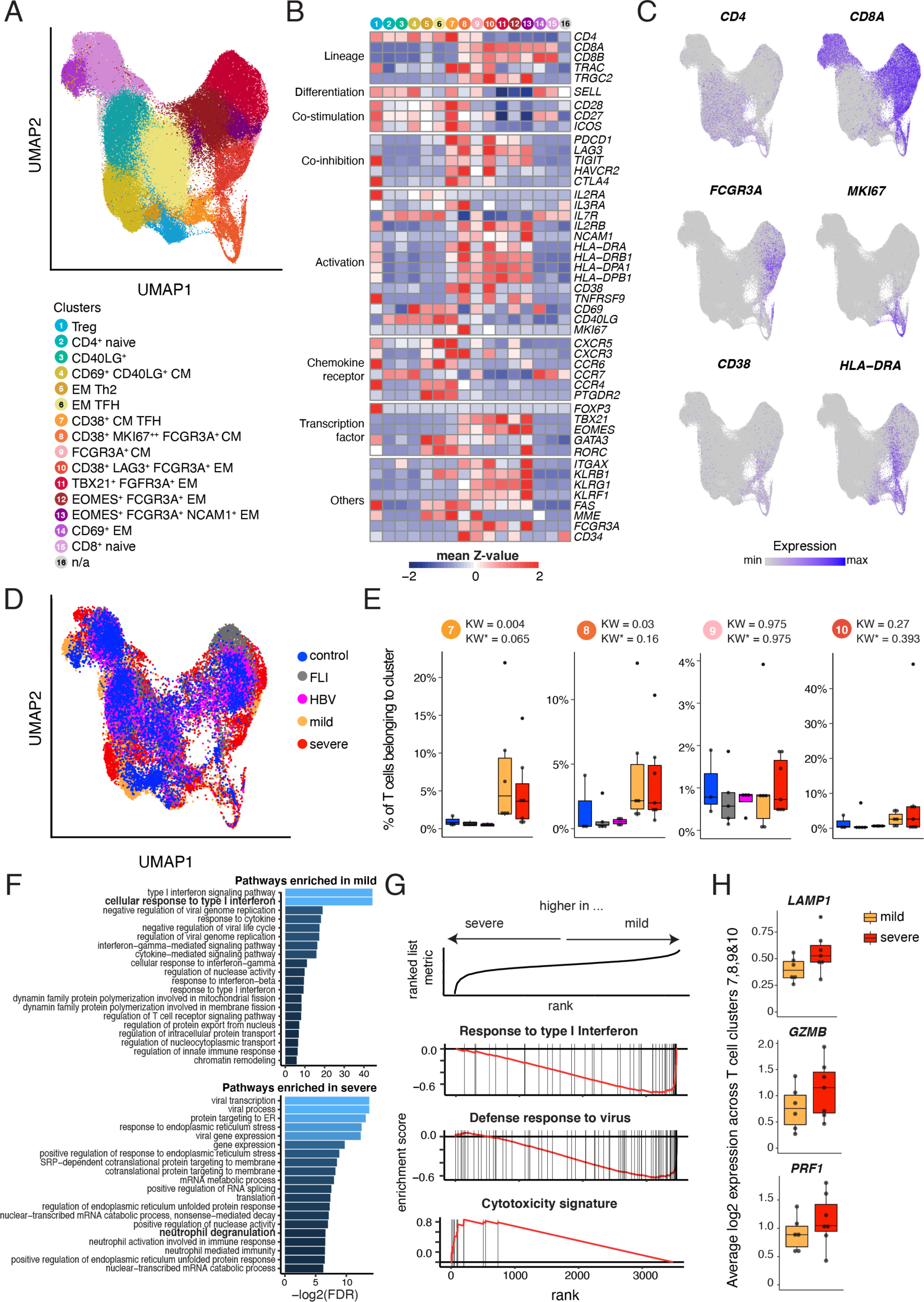
Single-cell transcriptomics of T cells during acute mild and severe COVID-19. **A**, UMAP of T cell clusters retrieved from merging scRNA-seq libraries of 5’-libraries of PBMC samples and 5’-libraries of enriched CD38^+^ CD4^+^ and CD8^+^ T cells collected from controls (n = 3), FLI (n = 5), HBV (n = 4), mild COVID-19 (n = 9) and severe COVID-19 (n = 12) patients. **B**, Heatmap of T cell cluster retrieved from merging scRNA-seq libraries of 5’-libraries of PBMC samples and 5’-libraries of enriched CD38^+^ CD4^+^ and CD8^+^ T cells collected from controls (n = 3), FLI (n = 5), HBV (n = 4), mild COVID-19 (n = 9) and severe COVID-19 (n = 12) patients. **C**, UMAPs of T cell clusters as shown in A with superimposed *CD4*, *CD8A*, *FCGR3A*, *MKI67*, *CD38* and *HLA-DRA* expression. **D**, UMAP of T cell clusters as shown in A with cells coloured according to disease group origin: blue, age-matched controls (n = 3); gray, FLI (n = 5); pink, HBV (n = 4); yellow, mild COVID-19 acute phase (n = 6); red, severe COVID-19 acute phase (n = 7). For visualization purposes, cells were downsampled to 5000 cells per disease group. **E**, Box plots of a selection of scRNA-seq T cell clusters whose abundances is either higher in both mild and severe COVID-19 (clusters 7 & 8) or which display *FCGR3A* expression and show a tendency to be increased in severe COVID-19 (clusters 9 & 10) generated from controls (n = 3), FLI (n = 5), HBV (n = 4), mild COVID-19 acute phase (n = 6) and severe COVID-19 acute phase (n = 7) patient samples. KW* shows the adjusted p-value (Benjamini-Hochberg) of a Kruskal-Wallis test. **F**. Bar plot indicating the negative log2-transformed adjusted p-value (Benjamini-Hochberg) of the 20 most significant enriched pathways that are (top) upregulated in mild COVID-19 acute phase, compared to severe COVID-19 acute phase, (bottom) upregulated in severe COVID-19 acute phase, compared to mild COVID-19 acute phase. Pseudobulk gene expression was calculated per sample among scRNA-seq T cell clusters 7, 8, 9 & 10. **G**, Enrichment plots from GSEA performed on the ranked gene list of the comparison severe vs. mild COVID-19. The graph shows the mapping of the signature genes on the ranked gene list. The curve corresponds to the running sum of the weighted enrichment score (ES). The ranked gene list was calculated from the normalized pseudobulk expression data of severe and mild COVID-19 acute phase among scRNA-seq T cell clusters 7, 8, 9 & 10. **H**, Box plots of the average log2-transformed expression among T cell clusters 7, 8, 9 & 10 from mild (n = 6) and severe (n = 7) COVID-19 acute samples, for three genes included in the cytotoxicity signature (*LAMP1*, *GZMB* and *PRF1*).

Thus, scRNA-seq analysis of samples from two cohorts strongly supported our CyTOF finding of a subset of activated CD16 expressing T cells across the major T cell compartments in severe COVID-19 and identified an increase in cytotoxicity-associated transcriptional programs.

### CD8^+^ T cells from severe COVID-19 patients display CD16-mediated degranulation potential leading to chemokine production by co-cultured primary lung microvascular endothelial cells

CyTOF and scRNA-seq analyses identified two main T cell activation features i) formation of highly activated, proliferating TFH-like CD4^+^ cells and CXCR3 expressing CTLs independent of disease severity, and ii) activated CD16 expressing T cells specific for severe COVID-19. Since TFH cells are known to promote B cell help (Crotty, 2014) and since we observed a trend for faster formation of activated TFH-like cells in severe COVID-19 patient samples, we tested whether SARS-CoV-2-specific antibody responses were more pronounced in those patients. Indeed, serum concentrations of SARS-CoV-2-specific IgA, but particular IgG levels were higher in severe COVID-19 (Figure 3A). These findings are in line with previous reports stating highest levels of both IgA and IgG in severely ill COVID-19 patients (Garcia-Beltran et al., 2021). Furthermore, maximal antibody levels determined during the second week post-symptom onset correlated positively with the cell proportions of the TFH-like CyTOF cluster 7 (Figure 3B), whereas the CD16^+^CD4^+^ T cell cluster 8 did not correlate with antibody levels (IgA: R^2^= 0.11, p= 0.19; IgG: R^2^= 0.18, p= 0.08; data not shown).

**Figure 3.**
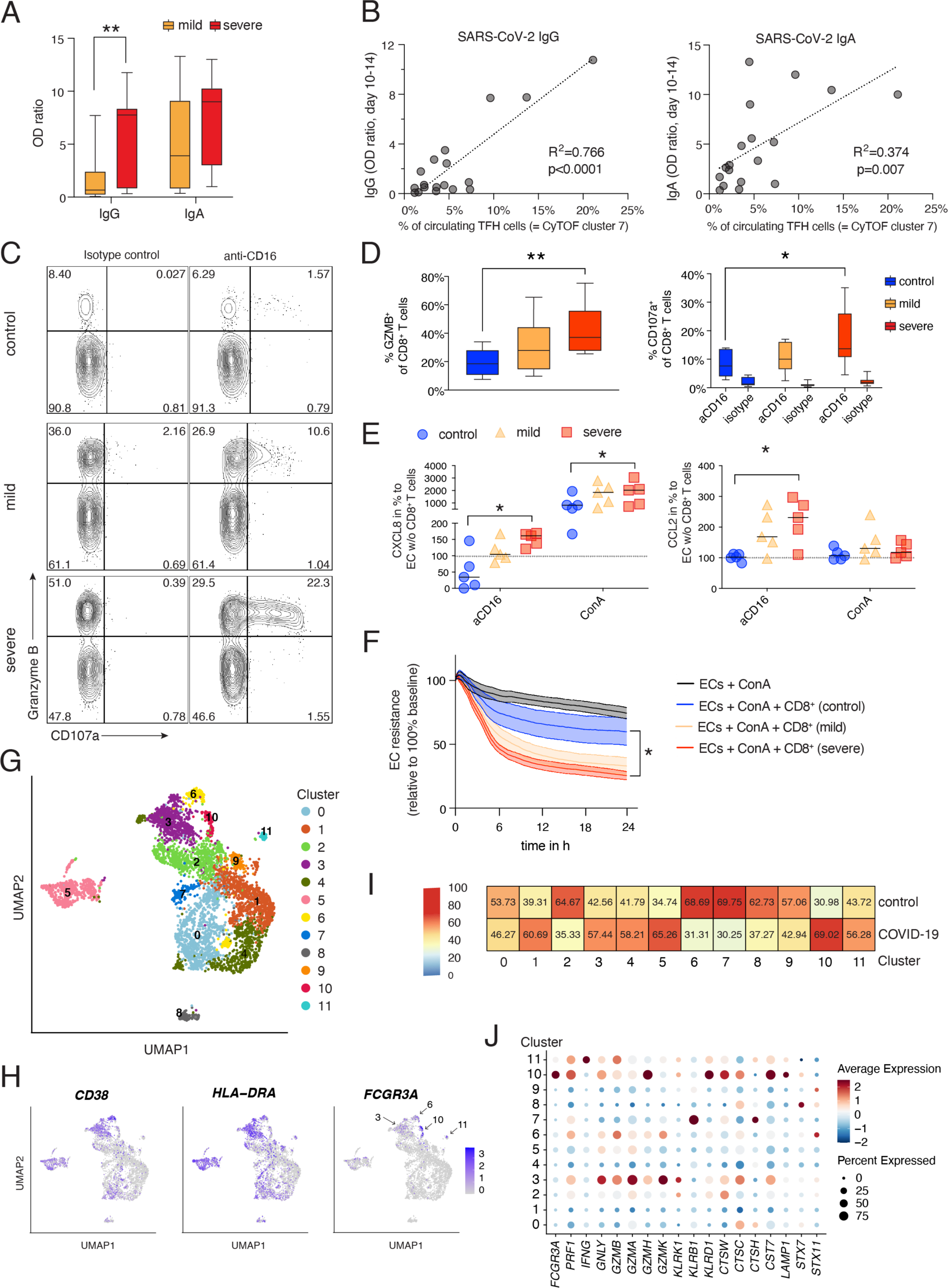
Increased degranulation and cytotoxic potential of T cells from severe COVID-19. **A**, Box and whisker (5–95 percentile) plots of SARS-CoV-2-specific IgG and IgA antibody levels detected as described in material and methods in serum samples from mild (n = 15) and severe (n = 17) COVID-19 patients collected between day 10 and 14 post symptom onset. Wilcoxon test **p < 0.01. **B**, Linear regression analysis of circulating TFH cell proportions (CyTOF T cell cluster 7) determined in whole blood samples collected from mild (n = 8) and severe (n = 11) COVID-19 from cohort 1 during day 5 and 14 post-symptom onset and SARS-CoV-2-specific IgG and IgA serum levels. **C**, Exemplary dot plots revealing the degranulation capacity of CD8^+^ T cells from PBMCs of a mild and severe COVID-19 patient as well as a non-infected control proband. Degranulation capacity is defined by the increase of cell surface CD107a expression upon stimulation with anti-CD16 antibody-coated or isotype-coated beads. **D**, Box and whisker (5–95 percentile) plots summarising the Granzyme B expression (unstimulated) and degranulation capacity of CD8^+^ T cells from PBMCs of mild (n = 9) and severe (n = 10) COVID-19 patients as well as non-infected controls (n = 6) defined by their increase of cell surface CD107a expression upon stimulation with anti-CD16 antibody-coated or isotype-coated beads. Wilcoxon test *p<0.05, **p < 0.01. **E**, Box and whisker (5–95 percentile) plots summarising the normalised release of CXCL8 and CCL2 by primary lung endothelial cells co-cultured with CD8^+^ T cells enriched from PBMCs of mild (n = 6) and severe (n = 5) COVID-19 patients as well as non-infected controls (n = 5) upon stimulation with ConA or anti-CD16 antibody-coated beads measured by multiplex assay. Wilcoxon test *p<0.05. **F**, Endothelial cell resistance upon stimulation with ConA alone (n = 5) or additional co-culture with CD8^+^ T cells enriched from PBMCs of mild (n = 6) and severe (n = 5) COVID-19 patients as well as non-infected controls (n = 5). Kruskal-Wallis test *p<0.05. **G**, UMAP of T cells generated from the BAL sample dataset of Wauters et al. (Wauters et al., 2021), including COVID-19 (n = 22) and control pneumonia (n = 13) samples. **H**, UMAPs of T cell clusters as shown in G with superimposed *CD38*, *HLA-DRA* and *FCGR3A* expression. **I**, Confusion matrix showing the percentage of the relative contribution of each group in each cluster. Each group was normalized to the same total number of cells to avoid biases derived from different cell numbers. **J**, Dot plot of the expression of the genes included in the “Cytotoxicity” signature as applied in the analyses of PBMC samples (Figure 2, S3) and *FCGR3A* in the T cell clusters as shown in G. The dots are colored by the scaled gene expression across the clusters and the size is proportional to the ratio of cells expressing the specific gene.

Next, we investigated the functional properties related to the CD16 expressing CD4^+^ and CD8^+^ T cell clusters. Since CD16 is known to mediate antibody-mediated degranulation of NK cells (Moretta et al., 2008), we tested whether T cells from severe COVID-19 patients display enhanced CD16-dependent degranulation potential. As a surrogate for immune complex-mediated T cell stimulation, we assessed intracellular Granzyme B and cell surface CD107a in PBMCs from mild or severe COVID-19 patients as well as controls after a 6-hour incubation with anti-CD16 antibody or isotype antibody--coated beads (Figure 3C, D). As predicted by scRNA-seq, samples from severe COVID-19 patients contained significantly more Granzyme B expressing CD8^+^ T cells compared to controls. Furthermore, stimulation with anti-CD16 but not isotype-coated beads elicited stronger degranulation (CD107a surface expression) of severe COVID-19 CD8^+^ T cells compared to T cells from mild COVID-19 (Figure 3D). In contrast, control T cells had very low degranulation potential.

One of the most likely implications of CD16 engagement in severe COVID-19 is enhanced T cell degranulation during interaction with endothelial cells (Bagnato and Harari, 2015; Degauque et al., 2021). Indeed, based on findings from autopsies in severe COVID-19, T cell infiltration along with endothelial cell injury known as lymphocytic endotheliitis has been observed (Miyazawa, 2020; Varga et al., 2020). One could envision that immune complexes made of virus particles and antibodies elicit degranulation of CD16 expressing T cells which contributes to endothelial injury. To test this hypothesis, we co-cultured primary lung microvascular endothelial cells with enriched non-naive (CD45RA^+/-^, CCR7^-^) CD8^+^ T cells from mild or severe COVID-19 as well as control T cells in the presence of anti-CD16 antibody. Subsequently, release of inflammatory mediators was analysed (Figure 3E). Anti-CD16-triggered severe COVID-19 T cells elicited enhanced CXCL8 (IL-8) and CCL2 (MCP-1) release by co-cultured endothelial cells. Importantly, chemokines were produced by endothelial cells as we did not observe chemokine release by anti-CD16-triggered T cells cultured alone (data not shown). We also analysed whether non-naive CD8^+^ T cells from mild and severe COVID-19 have a greater potential to amplify Concanavalin A (ConA)-mediated endothelial cell activation and loss of endothelial cell barrier function compared to control T cells. T cells from both mild and severe COVID-19 patients showed a higher potential to reduce transendothelial electrical resistance compared to control T cells, but this effect was only significant for T cells from severe COVID-19 (Figure 3F).

Next, we investigated whether CD16 expressing T cells are found locally in COVID-19. To study this we made use of a published scRNA-seq data set of bronchoalveolar lavage (BAL) samples from COVID-19 patients and patients with non-COVID-19 pneumonia (Wauters et al., 2021). Cluster analysis of the T cell space generated 11 clusters (Figure 3G). Whereas most T cell clusters contained *CD38* and *HLA-DRA* transcribing cells, *FCGR3A* expression was more or less confined to cluster 3, 6, 10 and 11 (Figure 3H). The abundance of cluster 3, 10 and 11 was higher in COVID-19 samples (Figure 3I). Importantly, cluster 10, showing the strongest enrichment in COVID-19 samples, was characterized by the highest *FCGR3A* expression. In addition, T cells belonging to this cluster transcribed nearly all genes of the cytotoxicity signature including *LAMP1* at the highest level (Figure 3J).

These findings support our hypothesis of a COVID-19-specific generation and local accumulation of CD16^+^, highly cytotoxic T cells, which can induce activation and injury of lung endothelial cells. T cell-induced release of chemoattractants CXCL8 and CCL2 can contribute to increased infiltration of neutrophils and monocytes in COVID-19 pneumonia.

### *FCGR3A* expressing T cell clones induced during acute severe COVID-19 persist and maintain their increased cytotoxic potential

Knowing that CD16 expressing T cells from severe COVID-19 patients display enhanced cytotoxic properties potentially contributing to organ damage, we analysed whether they persist (in circulation) after clearance of the acute infection. To do so, we obtained VDJ sequence information in addition to the gene expression data of T cells from acute and convalescent samples of mild and severe COVID-19 as well as FLI, HBV and controls, allowing us to study the fate of early expanded T cell clones during convalescence at months 3 to 4 post-symptom onset (Figure 4 A-E). First, we analysed whether the individual COVID-19 T cell clusters differ in their clonal enrichment. Interestingly, the *FCGR3A* expressing SARS-CoV-2-specific scRNA-seq T cell clusters 8 and 10 (introduced in Figure 2) showed a high level of clonal enrichment during acute COVID-19 infection, which was rather low for cluster 7, the cluster containing solely CD4^+^ TFH-like cells (Figure 4A). Consequently, only clones belonging to cluster 8, 9 and 10, composed of mixed *CD4*/*CD8A* or *CD8A*/*TCRgd* T cell clusters, displayed a high degree of persistence with up to 50% of the TCR clones being recovered in convalescent samples (Figure 4B). Furthermore, clones expanded in the highly proliferating cluster 8 showed a higher persistence in severe COVID-19 patients (Figure 4B). Following the clone-specific VDJ sequences also allowed us to track in which cluster these clones accumulated during convalescence, in other words, whether they either kept or changed their molecular phenotype as assessed by scRNA-seq (Figure 4C, D). While the T cells expanded during the acute infection and belonging to the SARS-CoV-2-specific clusters 7, 8 and 10 dropped dramatically during convalescence (Figure 4D) the respective T cell clones from clusters 8 and 10 as determined by TCR clonal fate analysis evolved into T cells mainly identified in clusters 11 and 12 which were still characterized by *FCGR3A* expression (Figure 4C, D). We therefore tested whether the convalescent clusters also maintained their enhanced cytotoxic potential (Figure 4E). Indeed, GSEA revealed increased expression of genes from the cytotoxicity signature among clusters 6, 11 and 12 T cells of COVID-19 patients versus controls, with an evident difference even when comparing severe versus mild COVID-19 (Figure 4E).

**Figure 4.**
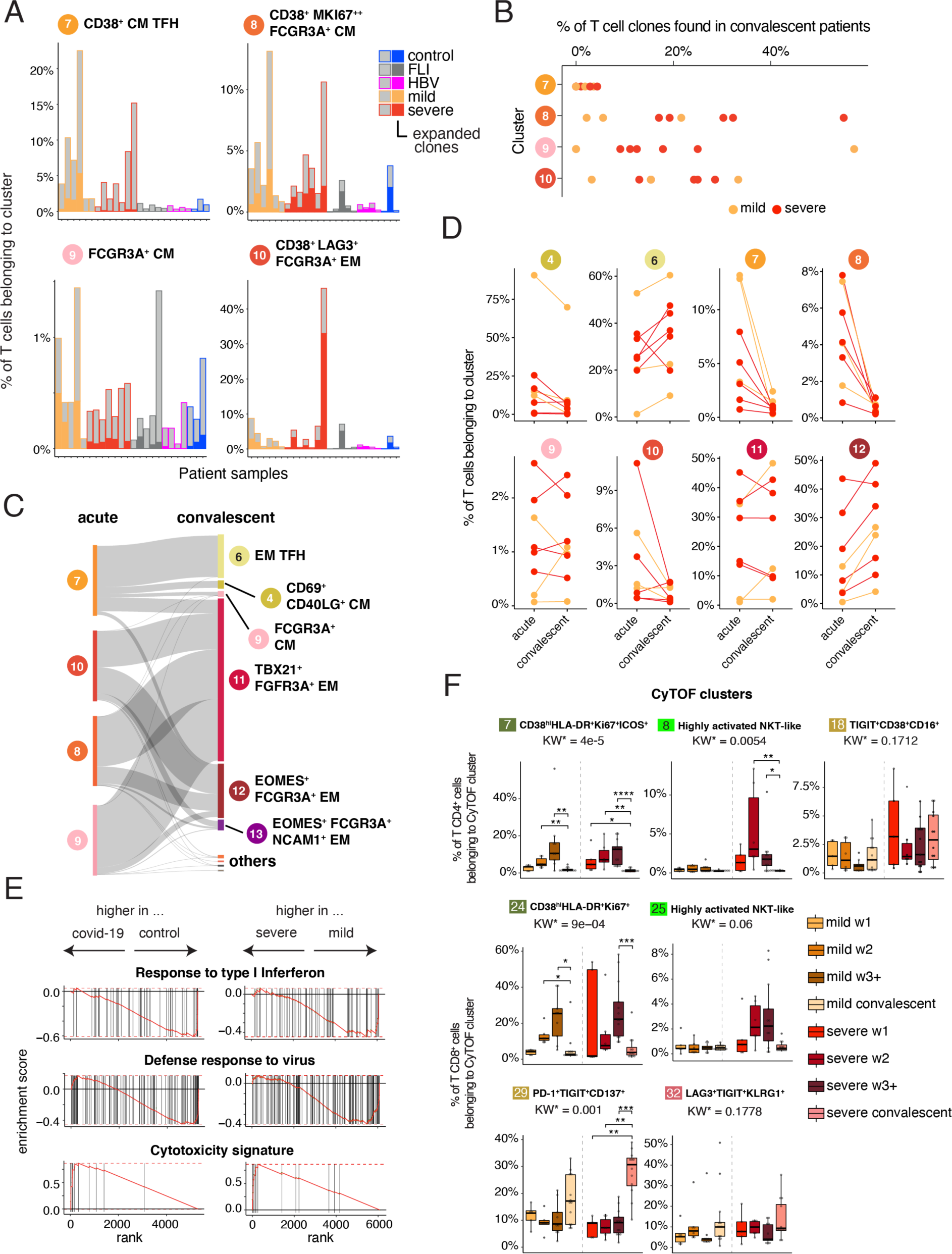
Time-dependent evolution and phenotype of T cell clones expanded during acute COVID-19. **A**, Percentage of expanded and non-expanded T cell clones in clusters 7, 8, 9 & 10. A cell that has the same clonotype in more than 1 per 1000 cells over all T cells per patient (controls, n = 3; FLI, n = 5; HBV, n = 4; mild COVID-19 acute phase, n = 6; severe COVID-19 acute phase, n = 7) was considered as an expanded clone. **B**., Percentage of T cell clones from clusters 7,8,9 & 10 acute phase found in convalescent samples (mild COVID-19, n = 3; severe COVID-19, n = 5) **C**, Flow diagram representing the cluster trajectory of clones present in acute (left) and convalescent (right) COVID-19 samples (mild COVID-19, n = 3; severe COVID-19, n = 5). **D**, Percentage of cells in selected clusters for each COVID-19 sample (mild COVID-19, n = 3; severe COVID-19, n = 5). **E**, Enrichment plots from GSEA performed on the ranked gene list of the comparison (left) control vs. convalescent COVID-19 (mild and severe), and (right) severe vs. mild convalescent COVID-19. The graph shows the mapping of the signature genes on the ranked gene list. The curve corresponds to the running sum of the weighted enrichment score (ES). The ranked gene list was calculated from the normalized pseudobulk expression data per sample among scRNA-seq T cell clusters 6,11 & 12. **F**, Box plots summarising the percentage of cells belonging to the indicated CD4^+^ (7, 8 & 18) and CD8^+^ (24, 25, 29 & 32) CyTOF cluster in samples from mild and severe patients during the acute (mild n = 20, severe n = 24) and convalescent phase (mild n = 13, severe n = 12). KW* shows the adjusted p-value (Benjamini-Hochberg) of a Kruskal-Wallis test. The abundance of each cluster was compared between severity groups via adjusted Dunn’s post-hoc test (Benjamini-Hochberg) for clusters with KW* < 0.05. All combinations were tested, only comparisons between the acute and convalescent phase within each COVID-19 severity group are shown (*p<0.05, **p < 0.01, ***p < 0.001, ****p < 0.0001).

Using the VDJ sequence information allowed us also to define whether the activated *FCGR3A*^+^ T cell clusters are mainly composed of NKT cells known to show innate immune cell functions (Krovi and Gapin, 2018). With the the vast majority of iNKT cells expressing an identical TCRα chain (*TRAV10-TRAJ18*) paired to a restricted set of TCRβ chains (*TRBV25*), we determined the proportion of *TRAV10-TRAJ18-TRBV25* pairing T cell clones across all T cell clusters. The median proportion of T cells expressing the TCR alpha-beta pair and thus resembling iNKT cells was 4.7%. Apart from cluster 10, we did not observe a major iNKT cell enrichment in the described COVID-19 T cell clusters (C7 = 3.73%, C8 = 6.52%, C9 = 2.12%, C10 = 13.46%). However, this shows that although the proportion of TRAV*10-TRAJ18-TRBV25* pairing T cell clones was higher in cluster 10, most clones expressed other alpha and beta chain genes and thus do not resemble iNKT cells.

Furthermore, we performed CyTOF analysis of samples during COVID-19 convalescence (Figure 4F). Similar to the scRNAseq findings, we observed alterations in T cell cluster abundances during convalescence. For CD8^+^ T cells, we observed a strong increase in T cell proportions for cluster 29, which belongs to the effector memory meta cluster and is characterized by PD-1, TIGIT as well as CD137 expression. In line with the scRNA-seq data for which we detected a decline of the acutely expanded T cell clusters, proportions of highly activated (clusters 7 and 24) and CD16 expressing NKT-like (clusters 8 and 25) CD4^+^ and CD8^+^ clusters decreased in the convalescent phase. However, CD16 expressing T cells did not disappear. In the CD4^+^ T cell compartment, we observed an increase in the CD16 expressing late differentiated TEMRA cluster 18, which was more pronounced in severe COVID-19 patients. Similarly, proportions of T cells belonging to the CD8^+^ CD16 expressing TEMRA cluster 32 were higher in the convalescent phase.

Thus, in summary, CD16 expressing CD8^+^ T cells in particular of severe COVID-19 patients persist during convalescence, adopt a more differentiated, CD62L-negative phenotype but maintain their high cytotoxic potential.

### Age-dependent reinforcement of C3a generation promotes differentiation of CD16 expressing highly cytotoxic T cells

We next elucidated the mechanisms leading to accumulation of activated CD16 expressing highly cytotoxic T cells in severe COVID-19.

Since mortality and severe morbidity in COVID-19 disproportionately affects those of advanced age (Nanda et al., 2020), we investigated whether formation of CD16 expressing T cells is associated with increased age. We utilized flow cytometry data available from our recently published control cohorts, in which we assessed age- and sex-dependent changes of peripheral blood immune cell populations in healthy individuals (Kverneland et al., 2016). Also blood samples from controls contain some CD16 expressing CD4^+^ and CD8^+^ T cells, yet at very low proportions (Figure 5A), which is in line with the here-reported findings (Figure 1+S1). Especially for CD8^+^ T cells, we detected significantly higher proportions of CD16 expressing cells in samples from older individuals, supporting our hypothesis of an age-dependent increase.

**Figure 5.**
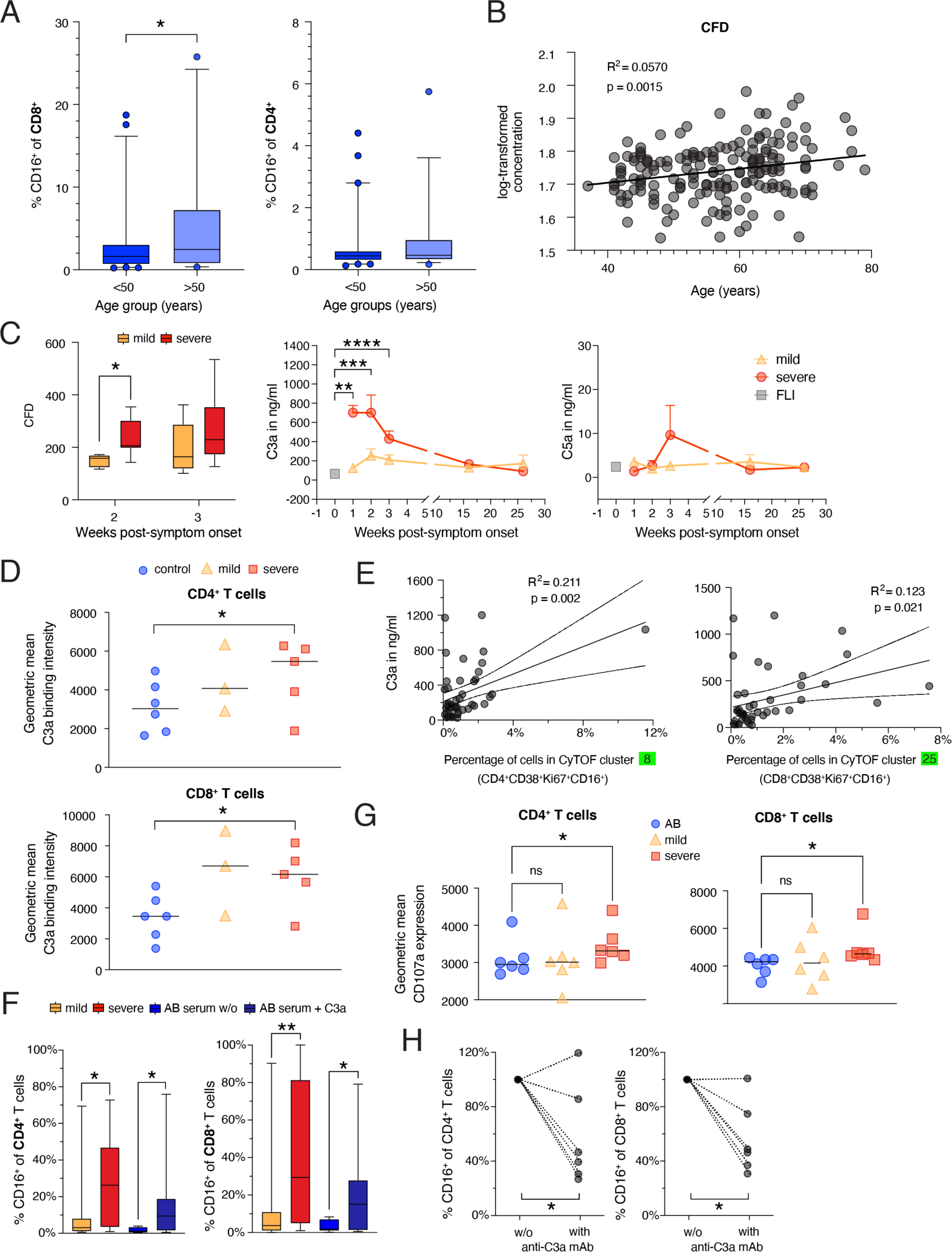
Age-dependent enforcement of C3a generation promotes differentiation of CD16 expressing highly cytotoxic T cells. **A**, Box and whisker (5–95 percentile) plots showing the proportions of CD16 expressing CD8^+^ and CD4^+^ T cells in whole blood samples of controls (n = 95) according to their age (<50 n = 59, >50 n = 36). Wilcoxon test *p<0.05. **B**, Linear regression analysis of age and plasma CFD levels determined by mass spectrometry in samples from the Generation Scotland study (Messner et al., 2020). **C**, Box and whisker (5–95 percentile) plots summarising the CFD plasma levels determined by mass spectrometry in samples collected from mild (week 2, n = 5; week 3, n = 7) and severe (week 2, n = 7; week 3, n = 17) COVID-19 patients during week two or three post-symptom onset. Wilcoxon test *p<0.05. Longitudinal changes of plasma C3a and C5a concentrations in samples from mild (n = 12) and severe (n = 17) COVID-19 and FLI (n = 8). Kruskal-Wallis and Dunn’s multiple comparison test **p < 0.01, ***p < 0.001, ****p < 0.0001. **D**, Scatter plots showing the differences in C3a binding capacity of non-naive CD4^+^ and CD8^+^ T cells enriched from PBMCs of mild (n = 3) or severe (n = 5) COVID-19 patients or controls (n = 6) determined by flow cytometry. Wilcoxon test *p<0.05. **E**, Linear regression analysis of the proportions of T cells belonging to CyTOF cluster 8 or 25 and plasma C3a levels in acute COVID-19 samples (n = 26). **F**, Box and whisker (5–95 percentile) plots showing the percentage of CD16 expressing CD4^+^ and CD8^+^ T cells upon one-week stimulation of enriched CD3^+^ T cells from controls with anti-CD3/ CD28 antibodies and IL-2 in medium containing serum from mild (n = 19), severe (n = 19) COVID-19 patients or AB serum in the presence or absence of recombinant C3a (n = 8). Wilcoxon test *p<0.05, **p<0.01. **G**, Scatter plots showing the cell surface CD107a expression level of CD4^+^ and CD8^+^ T cells upon one-week stimulation of enriched CD3^+^ T cells from controls with anti-CD3/CD28 antibodies and IL-2 in medium containing serum from mild (n = 6), severe (n = 6) COVID-19 patients or AB serum (n = 6). Friedmann and Dunn’s multiple comparison test *p<0.05. **H**, Scatter plots revealing the changes in the proportions of CD16 expressing CD4^+^ and CD8^+^ T cells upon one-week stimulation of enriched CD3^+^ T cells from controls with anti-CD3/CD28 antibodies and IL-2 in medium containing serum from mild or severe COVID-19 patients upon neutralisation of C3a (n = 6). Wilcoxon test *p<0.05.

Next, we sought to identify signals eliciting CD16 expression on T cells. A key feature of severe COVID-19 is increased complement generation and activation (Carvelli et al., 2020; Ma et al., 2021; Sinkovits et al., 2021). The complement system is an integral part of the innate immune defense involved in opsonization, chemotaxis and pathogen lysis (Noris and Remuzzi, 2013). Apart from these well-known complement activities, it has become clear that its effector functions also extend to an instruction of the adaptive immune system (Lubbers et al., 2017). In particular, complement enhances T cell activation as T cells are known to express receptors for C3a and C3b and react to triggering these receptors (Arbore et al., 2018; Hess and Kemper, 2016; West et al., 2018).

To corroborate a potential link between aging and an increase in complement activity, we analyzed the correlation of the complement component complement factor D (CFD) with age in the generation Scotland study (Figure 5B, (Messner et al., 2020)), which revealed a clear age-dependent increase of CFD. We then extended these findings to COVID-19 patients included in our CyTOF analysis and showed that particularly severe COVID-19 at early time points were characterized by significantly elevated CFD plasma concentrations (Figure 5C, left panel).

CFD is a serine protease that catalyses an important step in the formation of the active C3 convertase of the alternative pathway also known as spontaneous “tick-over” mechanism of complement activation (Noris and Remuzzi, 2013). Thus, higher CFD expression levels result in higher levels of C3a. Indeed, C3a plasma levels detected in severe COVID-19 patients during the first three weeks after symptom onset exceeded C3a levels detected in the plasma of mild COVID-19, or other acute respiratory infections (Figure 5C, middle panel). C5a levels peaked in weak three post onset of symptoms but did not reach significance (Figure 5C, right panel). We next assessed whether T cells show elevated binding potential for C3a in COVID-19. Both, CD4^+^ and CD8^+^ T cells from COVID-19 patients regardless of disease severity displayed higher C3a binding potential than cells from controls (Figure 5D). Finally, plasma C3a levels in COVID-19 patients measured at week 2 post-symptom onset correlated with proportions of COVID-19-specific activated CD16 expressing CD4^+^ and CD8^+^ CyTOF clusters (cluster 8+25) (Figure 5E).

To test whether there is a direct link between plasma / serum protein components and differentiation of activated CD16 expressing T cells, we stimulated enriched CD3^+^ cells from healthy unexposed controls with plate-bound anti-CD3/CD28 antibodies in the presence of recombinant IL-2 and serum from mild or severe COVID-19 patients or control AB serum. As shown in figure 5F, addition of serum from severe COVID-19 patients resulted in a 10- and 20-fold increase of CD16 expressing CD4^+^ and CD8^+^ T cells, respectively, which was significantly higher compared to the increase observed when adding serum from mild COVID-19 patients. Furthermore, addition of recombinant C3a to cells stimulated in the presence of control AB serum enhanced formation of CD16 expressing CD4^+^ and CD8^+^ T cells (Figure 5F). The *in vitro*-generated T cells did not only phenotypically resemble the ones identified in our CyTOF analyses but also shared functional properties as T cells stimulated in the presence of serum from severe COVID-19 patients displayed a higher degranulation potential (Figure 5G).

Finally, we tested whether C3a is responsible for the altered T cell differentiation potential of serum samples from severe COVID-19 patients. Indeed, neutralisation of C3a resulted in reduction of CD16^+^ T cells in most T cell differentiation cultures (Figure 5H).

In summary, complement split products such as C3a produced at high levels in severe COVID-19 generate an inflammatory milieu which promotes differentiation of CD16 expressing, highly cytotoxic T cells.

### Proportion of activated CD16^+^ T cells and complement protein expression determines outcome of COVID-19

Revealing a connection between the complement system and differentiation of activated CD16^+^ T cells in severe COVID-19, we investigated their relation to patient outcome in COVID-19.

We first analysed whether proportions of activated CD16 expressing T cells were higher in severe COVID-19 patients who died during follow-up in both study cohorts. In order to define an objective way to select the sum of all activated CD16^+^ TCRab^+^ clusters of our CyTOF data set (cohort 1), we computed an activation value for each cluster in the TCRab^+^ space, defined as the mean of the average z-scored expression of activation markers (CD25, HLA-DR, CD38, CD137, CD69, and Ki67). Then, clusters with an activation value higher than the average, and with an average z-scored CD16 expression higher than 1, were considered activated CD16^+^ TCRab^+^ T cell clusters (CyTOF cluster: C8 = CD4^+^ central memory highly activated NKT-like, C12 = CD4^+^ effector memory highly activated NKT-like, C25 = CD8^+^ central memory highly activated NKT-like). We observed a trend to higher percentages of activated CD16^+^ TCRab^+^ cells among all CD4^+^ and CD8^+^ T cells in samples from severe COVID-19 patients who deceased(non-survivor) compared to those who survived (survivor) (Figure 6A, second left panel). Furthermore, proportions of CD3^+^CD16^+^HLA-DR^+^ T cells as measured in cohort 2 by multicolor flow cytometry also showed a trend to be higher in severe COVID-19 patients who deceased during follow-up (Figure 6A, middle panel).

**Figure 6.**
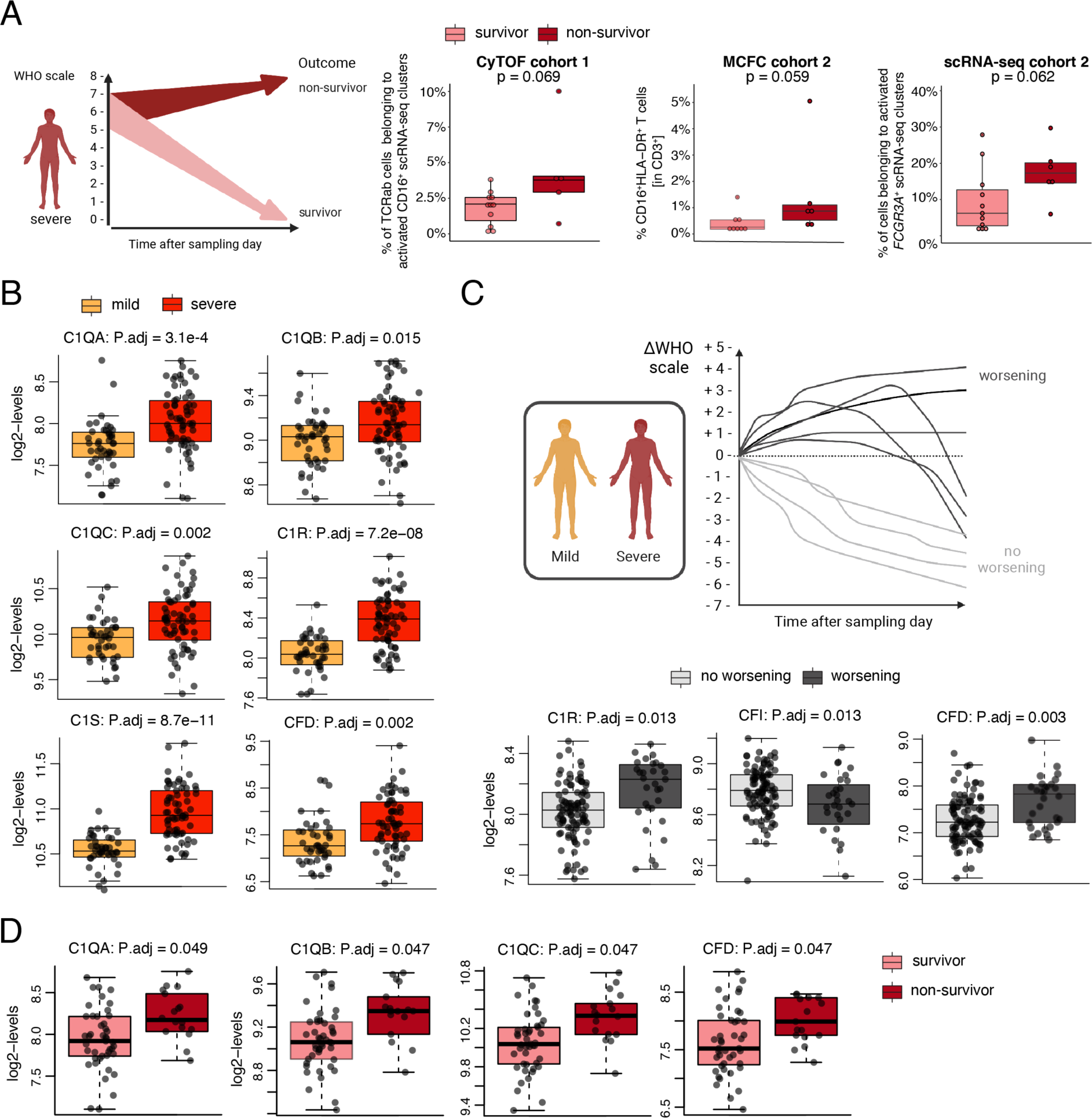
Proportion of activated CD16^+^ T cells and complement protein expression determines outcome in COVID-19. **A**, Subgroup analysis of activated CD16^+^ T cell proportions according to survival of severe COVID-19 patients. *Left panel*, cartoon summarizing the data analysis, data obtained from samples of severe COVID-19 patients (WHO 5-7) were divided according to patient survival. *Second left panel*, box plots of proportions of activated CD16^+^TCRab^+^ T cells across all T cell clusters determined by mass cytometry (whole blood, CyTOF cohort 1) generated from surviving (n = 11) and non-surviving (n = 5) week 2 or week 3+ severe acute COVID-19 patient samples. The sample with the highest WBC was selected per patient in case of repeated measurements. *Middle panel*, box plots of CD16^+^HLA-DR^+^ CD3^+^ T cells determined by flow cytometry (cohort 2) of week 2 or week 3+ samples from surviving (n = 8) or non-surviving (n = 6) severe COVID-19 patients collected during the acute infection. The sample with the highest WBC was selected per patient in case of repeated measurements. *Right panel*, proportions of activated *FCGR3A^+^* T cells (from scRNAseq clusters 8, 10 & 13, cohort 2) within the whole TCRab^+^ T cell space. The proportions of T cells between samples from surviving and non-surviving severe COVID-19 patients were compared using the Wilcoxon test. **B,** Box plots showing plasma expression levels of complement proteins upstream of C3a generation determined by mass spectrometry (proteomics) as described in material and methods which are significantly different between samples from mild (n = 44) and severe (n = 66) COVID-19 patients of cohort 1. The complement protein expression levels between samples from mild and severe COVID-19 patients were compared using the Wilcoxon followed by post-hoc Benjamini Hochberg test. **C,** Subgroup analysis of plasma complement protein expression levels upstream of C3a generation according to worsening of WHO scale. *Upper panel*, cartoon summarizing the data analysis, data obtained from samples of mild and severe COVID-19 samples were divided according to the subsequent WHO scale development (delta WHO). *Lower panel*, box plots showing plasma expression levels of complement proteins which are significantly different between samples from patients with non-worsening WHO scale (n = 91) and worsening WHO scale (n = 19) of mild or severe COVID-19 patients from cohort 1. The complement protein expression levels between the groups were compared using the Wilcoxon followed by post-hoc Benjamini Hochberg test. **D,** Subgroup analysis of plasma complement expression levels according to survival of severe COVID-19 patients. Box plots showing plasma expression levels of complement proteins upstream of C3a generation which are significantly different between samples from severe COVID-19 patients (WHO 5-7) who were divided according to patient survival (survivors, n = 48; non-survivors, n = 18). The complement protein expression levels between the groups were compared using the Wilcoxon followed by post-hoc Benjamini Hochberg test.

In addition, we tested whether the proportion of activated *FCGR3A* expressing scRNAseq clusters correlates with survival of severe COVID-19 patients. We applied a similar two-step strategy to select all activated *FCGR3A*^+^ TCRab^+^ clusters as described for the CyTOF data. All T cell clusters which showed an above mean activation value, calculated as mean z-score of activation markers as defined in Figure S3B, and clusters which showed *FCGR3A* transcription were selected (Figure S3B, cluster 10, 13 and 18). Again, the percentage of T cells belonging to activated *FCGR3A*^+^ T cell clusters showed a trend to be higher in severe COVID-19 patients who deceased (Figure 6A, right panel). Together, this difference was significant, when analysed across methods (CyTOF, MCFC, scRNAseq) and cohorts (p=0.011, Fisher’s method, df=6)

Next, we tested in a larger cohort whether expression of complement proteins upstream of C3a generation (C1QA, C1QB, C1QC, C1R, C1S, C2, C3, C4, C4A, C4B, C4BPA. C4BPB, CFB, CFD, CFH, CFI, MASP1, MASP2, MBL2) is associated with patient disease course and outcome. As shown in figure 6B, expression levels of positive regulators of the classical and alternative pathway such as C1QA, C1QB, C1QC, C1R, and CFD were higher in plasma samples of severe compared to mild COVID-19 patients (Figure 6B). We also analysed the complement protein expression level according to subsequent worsening of disease severity, i.e., increase of WHO scale. This also revealed higher expression of C1R and CFD in samples from patients who showed an increase in WHO scale during follow-up, whereas the expression of CFI, which inhibits the classical and lectin-dependent complement pathway was lower in samples from patients with subsequent disease progression (Figure 6C). Finally, amounts of C1QA, C1QB, C1QC and CFD were not only higher in severe COVID-19 but also determined their survival (Figure 6D).

Altogether these data further support the pathological role of complement-mediated formation of activated CD16^+^ T cells during severe COVID-19.

## Discussion

T cells play an important role in the control of acute respiratory infections. However, excessive T cell activation and altered phenotypes can contribute to infection-associated organ damage. Early after the first reports on immune profiles of COVID-19 patients (Sette and Crotty, 2021) discussions on their putative role in immune protection versus pathology started. Although evidence for an association of increased T cell activation with disease severity during the acute phase was emerging (Feng et al., 2020; Mathew et al., 2020; Peng et al., 2020; Thieme et al., 2020), functional consequences and causal mechanisms remain poorly studied. In our study, we provide evidence that SARS-CoV-2 infection - in contrast to other acute and chronic infections - promotes formation of highly activated and proliferating HLA-DR^+^CD38^hi^CD137^+^CD69^+^ T helper cells and CD8^+^ T cells independent of disease severity, although this response occurred faster in severe COVID-19 patients. More importantly, primarily in severe COVID-19 patients, we detected differentiation of activated CD16^+^ T cells, which showed an increased immune complex-mediated cytotoxic potential causing activation of lung microvascular endothelial cells. Expanded clones within the CD16^+^ T cell compartment persisted and maintained their high cytotoxic potential. Furthermore, our functional investigations identified complement factors and especially C3a as a driving signal for the differentiation of the altered activated T cell phenotype. In addition, proportions of activated CD16^+^ T cells and plasma complement protein expression levels determined outcome of severe COVID-19 patients. Thus, interaction of age-related increased production of complement components and SARS-CoV-2-triggered complement activation creates an inflammatory milieu which drives differentiation of T cells with high immunopathogenic potential.

A balanced T cell activation regarding magnitude and functional properties is decisive for the course of infection. Formation of CD8^+^ tissue-resident memory T cells (Trm) during primary infection is known to play an important protective role to restrain viral spread upon secondary influenza infections. Yet, enhanced accumulation of Trm cells in an imbalanced environment such as during aging can support excessive inflammation leading to organ damage and impaired repair (Goplen et al., 2020). In COVID-19, large numbers of such Trm-like CD8^+^ T cells have been identified in the airways (Liao et al., 2020).

Blood samples acquired during the acute phase of severe COVID-19, contain high numbers of HLA-DR^+^CD38^hi^Ki67^+^ in both CD4^+^ and CD8^+^ T cell compartments (Mathew et al., 2020; Rydyznski Moderbacher et al., 2020; Stephenson et al., 2021), a finding that we corroborated by CyTOF analysis. Most intriguing, we identified an elevated and activated T cell population expressing CD16 across the three major T cell compartments, namely CD4^+^, CD8^+^ TCRab^+^, and TCRgd^+^ T cells. These T cells show increased TCR-independent pathogenic potential. Thus, in severe COVID-19 T cell activation and functionality is altered due to an imbalanced, aged, and C3a-rich environment.

The activated CD16^+^ CD4^+^ and CD8^+^ T cells enriched in severe COVID-19 expressed high levels of chemokine receptors such as CXCR3 and CCR6 (Figure 1D). This clearly distinguished them from the other CD16^lo^ T cell clusters e.g. cluster 18 and promoting migration into the lungs (Oja et al., 2018; Shanmugasundaram et al., 2020). Indeed, BAL samples from COVD-19 compared to control pneumonia showed an enrichment of *FCGR3A^+^* T cells with high cytotoxic and degranulation potential (Figure 4G-J).

Whether the activated CD16 expressing CD4^+^ and CD8^+^ T cells are SARS-CoV-2 antigen specific remains unclear. At least for the *ex vivo*-determined HLA-DR^+^CD38^hi^Ki67^+^CD8^+^ T cells, a positive correlation with SARS-CoV-2-specific CD8^+^ T cells has been described (Rydyznski Moderbacher et al., 2020). However, the generation of CD16 expressing T cells can be also driven by bystander activation and/or homeostatic proliferation (Mathew et al., 2020). Therefore, antigen specificity for the here-described pathogenic T cells might not play a dominant role, as these cells elicited their excessive cytotoxic properties upon triggering of CD16 rather than peptide recognition.

In line with our findings, a very high proportion of T cells from acute COVID-19 patients express cytotoxic molecules such as Perforin and Granzyme B, with samples of severe patients displaying the highest expression levels (Shuwa et al., 2021). The increased cytotoxic profile persisted for up to six months and was associated with poorer recovery. CD16^+^ T cells identified here in severe COVID-19 did not only express higher levels of *PRF1* and *GZMB* but also *LAMP1* and *STX11* (Figure 2H, Figure S2B), which explains their increased general degranulation potential (Figure 3C,D) (Spessott et al., 2017). So far, CD16^+^ T cells with CD16-dependent cytotoxic properties have been described mainly in patients with chronic infections or inflammation (Björkström et al., 2008; Clémenceau et al., 2011; Jacquemont et al., 2020). In these conditions, CD16^+^ T cells displayed a more differentiated phenotype like the one adopted during COVID-19 convalescence (Figure 4C, D & F). Formation of activated CD16^+^ T cells during acute infections has not been described before. Furthermore, we are the first to identify upstream signals leading to CD16 expression on T cells. In acute COVID-19, we also identified elevated transcription of various granzyme genes including *Granzyme K* (Figure S2B). Interestingly, increased numbers of Granzyme K expressing effector memory T cells have been observed in blood samples of older individuals and these T cells were shown to augment cytokine and chemokine production by fibroblasts (Mogilenko et al., 2021). Furthermore, it was shown that extracellular Granzyme K proteolytically activates Protease-activated receptor-1 leading to increased release of IL-6 and CCL2 (MCP-1) by endothelial cells (Sharma et al., 2016). Particularly in severe COVID-19, we demonstrated that T cells also induce CCL2 (MCP-1) by co-culture with primary lung endothelial cells upon anti-CD16 mAb-mediated degranulation (Figure 3E). We also observed a higher production of CXCL8 by endothelial cells co-cultured with anti-CD16-stimulated CD8^+^ T cells from severe COVID-19 (Figure 3E). This establishes a general link between immune complex triggering of local CD16^+^ T cells and endothelial cell-mediated release of monocyte and neutrophil chemoattractants, a hallmark of severe COVID-19 (Rendeiro et al., 2021). While CD16^+^ T cells had been described before, they have not been linked to an acute infection such as severe COVID-19 and potential upstream signals inducing this molecular phenotype remained unclear. Notably, patients suffering from severe COVID-19 have been reported to display high levels of spike-reactive IgG with significantly reduced Fc fucosylation. This change in the Fc glycosylation pattern increases binding affinity to CD16, leading to increased CD16-mediated effector function and degranulation (Ferrara et al., 2011; Vivier et al., 2008; Willianne et al., 2021). As such, the distinct serological profile observed in severe COVID-19 with afucosylated, spike-directed IgG, and an inherently increased inflammatory capacity could further enhance the pathogenic potential of CD16 expressing T cells.

In a search for important environmental signals driving differentiation of activated CD16^+^ T cells, we detected a positive correlation between high serum C3a levels and proportions of CD16^+^ T cell clusters (Figure 5E). In line with our findings, it has been reported that serum C3 hyperactivation is a risk factor for COVID-19 mortality (Sinkovits et al., 2021), yet widespread complement activation and generation of C3a has also been described locally in severe COVID-19 patients (Gao et al., 2020). Disease-promoting activity of the complement system was observed for other corona viruses, as SARS-CoV infection caused less systemic inflammatory response and lung injury in C3 knock-out as compared to wild type mice (Gralinski et al., 2018). Furthermore, it has been shown that SARS-CoV-2 infection of lung epithelial cells induces transcription of complement genes leading to generation of activated C3a (Yan et al., 2021). Indeed, signalling via complement receptors such as C3AR1 and cell-autonomous complosome in human T cells enhances induction of CD4^+^ Th1 responses and cytotoxic function of CD8^+^ T cells (Arbore et al., 2018). Here, we show that increased C3a generation in severe COVID-19 patients promotes differentiation of CD16 expressing, highly cytotoxic CD4^+^ and CD8^+^ T cells (Figure 5F-H).

Interestingly, targeting distal complement effects by receptor blockade in a humanized preclinical model of SARS-CoV-2 infection prevented acute lung injury (Carvelli et al., 2020). Results from first clinical trials on complement inhibition in COVID-19 also showed promising effects resulting in reduced inflammation and faster normalization of neutrophil and lymphocyte counts (Mastellos et al., 2020; Polycarpou et al., 2020). In this context, C3 inhibition enabled a broader and better therapeutic potential as compared to C5 neutralisation (Mastellos et al., 2020).

### Limitations of the study and future directions

The relatively low number of matched acute and convalescent samples for scRNA-seq analysis limited the comparisons regarding differences in clonal persistence and phenotype between mild and severe COVID-19 patients. Thus, larger studies are needed to confirm the observed increased persistence of late differentiated, CD16 expressing, highly cytotoxic CTLs upon an acute severe disease course. Also, the limited number of convalescent samples did not allow us to perform correlations with patient recovery. Here, our major focus was to reveal immunopathogenic functions of severity-associated T cell populations during acute COVID-19 and to identify driving signals.

In this context, it will be of great interest to see whether application of the C3 inhibitor AMY-101 in COVID-19 patients with ARDS will ameliorate differentiation of CD16 expressing, cytotoxic T cells and thus endothelial cell injury and lymphocytic endotheliitis.

Taken together, particularly severe COVID-19 leads to an elevation of activated CD16^+^ T cells that link the elevated complement cascade via TCR-independent cytotoxic T cell functionality to endothelial damage and patient survival, thereby establishing a novel immunopathological link between the innate immune system, the adaptive immune compartment, and endothelial injury, which might constitute an important molecular axis explaining the vast spectrum of organ damage observed in COVID-19.

## STAR Methods

### Resource availability

#### Lead Contact

Further information and requests for resources and reagents should be directed to the Lead Contact, Birgit Sawitzki

#### Materials Availability

This study did not generate new unique reagents.

#### Data and Code Availability

Raw count data of the scRNA-seq experiments are deposited at GEO under the accession number GSE175450.

#### Study subject details

Samples from patients with COVID-19 were collected within two cohort studies (Schulte-Schrepping et al., 2020) designed to allow deep molecular and immunological studies of COVID-19 in blood. This study was primarily designed to describe immunological deviations in COVID-19 patients without intention of the development of new treatments or new diagnostics, and therefore sample size estimation was not included in the original study design.

#### Cohort 1 / Berlin cohort

This study includes a subset of patients enrolled between March 2 and April 30 2021 in the Pa-COVID-19 study, a prospective observational cohort study assessing pathophysiology and clinical characteristics of patients with COVID-19 at Charité Universitätsmedizin Berlin (Kurth et al., 2020). The study was approved by the Institutional Review board of Charité (EA2/066/20). Written informed consent was provided by all patients or legal representatives for participation in the study. The patient population included in all analyses of cohort 1 consists of 43 control donors (samples collected in 2019 before SARS-CoV-2 outbreak or collected in the fall of 2020 who did not have a SARS-CoV-2 infection), 8 patients presenting with flu-like illness but tested SARS-CoV-2-negative, 6 patients with chronic HIV infection, 5 patients with chronic HBV infection as well as 35 mild and 42 severe COVID-19 patients during the acute or convalescent phase (Figures 1A+B; Table S1). Information on age, sex, medication, and comorbidities is listed in Table S1. All COVID-19 patients were tested positive for SARS-CoV-2 RNA in nasopharyngeal swabs and allocated to mild (WHO 2-4, mild+moderate) or severe (WHO 5-8, severe+critical) disease according to the WHO clinical ordinal scale (https://www.who.int/blueprint/priority-diseases/key-action/COVID-19_Treatment_Trial_Design_Master_Protocol_synopsis_Final_18022020.pdf).

#### Cohort 2 / Bonn cohort

This study was approved by the Institutional Review board of the University Hospital Bonn (073/19 and 134/20). After providing written informed consent, 14 control donors and 22 COVID-19 patients were included in the study. In patients who were not able to consent at the time of study enrollment, consent was obtained after recovery. Information on age, sex, medication, and comorbidities are provided in Table S1. COVID-19 patients who tested positive for SARS-CoV-2 RNA in nasopharyngeal swabs were recruited at the Medical Clinic I of the University Hospital Bonn between March 30 and June 17, 2020 and allocated to mild (WHO 2-4, mild+moderate, n = 9) or severe (WHO 5-8, severe+critical, n = 13) disease according to the WHO clinical ordinal scale. Controls in cohort 2 were collected from healthy people or from otherwise hospitalized patients with a wide range of diseases and comorbidities including chronic inflammatory immune responses. These individuals were either tested negative for SARS-CoV-2, serologically negative or had no indication for acute COVID-19 disease based on clinical or laboratory parameters.

#### Antibodies used for mass cytometry (cohort 1)

All anti-human antibodies pre-conjugated to metal isotopes were obtained from Fluidigm Corporation (San Francisco, USA). All remaining antibodies were obtained from the indicated companies as purified antibodies and in-house conjugation was done using the MaxPar X8 labeling kit (Fluidigm, USA). Table S2 shows a detailed list of all antibodies used.

#### Sample processing, antigen staining and data analysis of mass cytometry-based immune cell profiling (cohort 1)

Sample processing, cell staining and acquisition was done as previously described (Schulte-Schrepping et al., 2020).

T cells were identified based on expression of CD3 and CD45, but exclusion of CD19^+^ and CD15^+^ cells. Pregating on CD8^-^TCRgd^-^, CD8^+^TCRgd^-^ and TCRgd^+^ cells was used to define CD4^+^, CD8^+^ and TCRgd^+^ T cells. Each T cell compartment was then clustered based on the expression of 30 markers: CD62L, CD45RO, CD28, CD27, CD226, ICOS, PD-1, LAG3, TIGIT, CD96, CD25, CD56, HLA-DR, CD38, CD137, CD69, Ki67, CXCR3, CXCR5, CCR6, CRTH2, KLRB1, KLRG1, KLRF1, CD95, CD10, CD16, CD34, CD123, CD11c, CD21, CD14, IgM, and IgD The batch-normalized CyTOF values (described in (Schulte-Schrepping et al., 2020)) were first transformed with the inverse hyperbolic sine function (asinh) and then z-score normalised per marker across all samples. We then clustered each T cell compartment excluding convalescent COVID-19 samples using Phenograph (Levine et al., 2015) with 30 nearest neighbours (k = 30). We originally found 50 clusters, which were annotated based on the average z-score transformed CyTOF expression of the markers in each cluster. Six clusters were merged in pairs, as each pair differed in one marker (KLRB1, CD11c, and CD21, respectively) resulting in 47 clusters. Cells from the convalescent samples were classified into the clusters previously found, via k-nearest neighbour (in Euclidean distance) using the knn function from “class” R package. (Figure S4). UMAPs were calculated across all acute and convalescent samples with the markers mentioned above excluding CD21, CD14, IgM, and IgD, using the R package “uwot” (arXiv:1802.03426 [stat.ML]), n_neighbors = 30, spread = 1, min_dist = 0.5, based on Euclidean distance).

The frequency of each cluster was calculated as the percentage of cells in each cluster for each patient and for each T cell compartment. Statistical testing for the difference in the frequency of each cluster was calculated with the adjusted Dunn’s post-hoc test (Benjamini-Hochberg) for clusters with significant Kruskal-Wallis test (adjusted p-value (Benjamini-Hochberg) < 0.05). For the non-weekly analysis, we considered the sample with the highest white blood cells (WBC) counts per patient. Similarly, for the weekly analysis, only the sample with highest WBC counts per week was included, and the repeated samples in the same week were excluded from the analysis.

### Blood processing and data analysis for multi-color flow cytometry

#### COVID-19 cohort (cohort 2)

Whole blood was prepared by treatment of 1ml peripheral blood with 10ml of RBC lysis buffer (Biolegend, USA). After one wash in DPBS, cells were directly processed for scRNA-seq (BD Rhapsody) or multi-color flow cytometry (MCFC). After RBC lysis, cells were washed with DPBS and from each sample 1-2 million cells were stained for flow cytometric analysis. Antibody staining was performed in DPBS with the addition of BD Horizon Brilliant Stain Buffer (Becton Dickinson, USA) for 30min at 4°C as described before (Schulte-Schrepping et al., 2020). To remove dead cells from the analysis a staining for dead cells was included (LIVE/DEAD Fixable Yellow Dead Cell Stain Kit; 1:1000 – Thermo Scientific, USA). To prevent any possible contamination of the operator, after staining, the samples were fixed for 5 minutes with 4% PFA. Flow cytometry analysis was performed on a BD Symphony instrument (Becton Dickinson, USA) configured with 5 lasers (UV, violet, blue, yellow-green, red). Flow cytometry data analysis was performed with FlowJo V10.7.1, CD16/HLA-DR double positive cells were gated from the total T lymphocytes (living/CD45^+^/ CD66b^-^/CD19^-^/CD3^+^).

#### Age-dependent control cohorts

We used our flow cytometry data sets (fcs files) described previously (Kverneland et al., 2016) to report the proportions of CD16 expressing CD4+ and CD8+ T cells. Briefly, whole blood samples collected from healthy controls spanning and age range between 20 and 84 years with equal distribution of females and males in each 10-year age bin were stained using the ONE Study antibody panels as previously described (Sawitzki et al., 2020; Streitz et al., 2013) and acquired on a 10-Color Navios Flow Cytometer (Beckman Coulter, USA). For the here reported results data from panel 1 were used. The flow cytometry data analysis was performed in Kaluza version 1.2 (Beckman Coulter, USA).

### Isolation of blood cells for scRNA-seq (cohort 1)

Human peripheral blood mononuclear cells (PBMCs) were isolated from heparinized whole blood by density gradient centrifugation over Pancoll (density: 1,077g /ml, PAN-Biotech, Germany). Subsequently the cells were counted, frozen and stored in liquid nitrogen. On the day of the experiment the frozen PBMCs were thawed in pre-warmed thawing medium (RPMI 1640, Gibco; 2% FCS, Sigma; 0,01% Pierce Universal Nuclease, Thermo Fisher, USA).

### 10x Genomics Chromium single-cell RNA-seq (cohort 1)

Approximately 2-3 x10^5^ PBMCs were resuspended in staining buffer (DPBS, Gibco; 0,5% BSA, Miltenyi Biotec, Germany; 2 mM EDTA, Gibco, Thermo Fisher Scientific, USA) and hashtagged with 0.5 µg Total-Seq-C™ Hashtag antibodies for 30 min at 4°C. After the incubation PBMCs were washed three times. Subsequently PBMC were counted and up to four different samples were pooled equally. The PBMCs were washed three times, resuspended in DPBS, filtered through a 40 µm mesh (Flowmi™ Cell Strainer, Merck, Germany) and counted using the C-Chip hemocytometer (NanoEntek, South Korea). The cell suspension was super-loaded with 40000 - 50,000 cells per lane, in the Chromium™ Controller for partitioning single-cells into nanoliter-scale Gel Bead-In-Emulsions (GEMs).

Remaining PBMCs were subjected to flow-cytometric sorting based on DAPI, CD3 (clone UCHT1), CD4 (clone RPA-T4), CD8 (RPA-T8) and CD38 (clone HB7) antibody staining and simultaneously hashtagged as described above. Live (DAPI-) CD3^+^CD4^+^CD38^+^ as well as CD3^+^CD8^+^CD38^+^ cells were sorted using the FACS Aria II (BD Biosciences, USA). Afterwards CD4^+^CD38^+^ and CD8^+^ CD38^+^ T cells from each donor were pooled equally and CD38^+^ T cells from up to four samples were pooled to equal proportions. The resulting cell pool was resuspended in DPBS, filtered through a 40 µm mesh (Flowmi™ Cell Strainer, Merck, Germany) and counted using the C-Chip hemocytometer (NanoEntek, South Korea). The cell suspension was super-loaded with 40,000 - 50,000 cells per lane, in the Chromium™ Controller for partitioning single-cells into nanoliter-scale Gel Bead-In-Emulsions (GEMs).

The Chromium Next GEM Single Cell 5′ Kit v1.1 was used for reverse transcription, cDNA amplification and library construction of the gene expression libraries (10x Genomics, USA). For additional VDJ and hashtag libraries the Chromium Single Cell V(D)J Enrichment Kit, Human T Cell (10x Genomics, USA) and the Chromium Single Cell 5’ Feature Barcode Library Kit (10x Genomics, USA) were used respectively. All libraries were prepared following the detailed protocols provided by 10x Genomics, quantified by Qubit Flex Fluorometer (Thermo Fisher, USA) and quality was checked using 2100 Bioanalyzer with High Sensitivity DNA kit (Agilent, USA). Sequencing was performed in paired-end mode with a S1 and S2 flow cell using NovaSeq 6000 sequencer (Illumina, USA).

### BD Rhapsody single-cell RNA-seq (cohort 2)

For the analysis of the T lymphocytes compartment of COVID-19 patients, the PBMC dataset from Schulte-Schrepping et al. (Schulte-Schrepping et al., 2020) was used. The dataset was provided as a Seurat object of the PBMCs samples from the original study including all pre-processing and filtering criteria as described in the original manuscript. **The data can be downloaded from FASTGenomics** [https://beta.fastgenomics.org/p/schulte-schrepping_covid19] **or directly explored using the web platform.**

### Data pre-processing of 10x Genomics Chromium scRNA-seq data (cohort 1)

The Cell Ranger Software Suite (Version 5.0.0) was used to process raw sequencing data, using the cellranger multi workflow and the GRCh38 references for gene expression and VDJ data and totalSeq antibody barcode sequences for feature barcoding data. Multiplexed samples from multiple donors were demultiplexed by detecting SNPs using CellSNP-lite (Huang and Huang, 2021).

### ScRNA-seq data analysis of 10x Chromium data (cohort 1)

#### Data quality control

For each sample, the number of RNA counts per cell was mean-normalized, and cells with a mean-normalized RNA count lower than 0.45 and higher than 5 were excluded from the analysis, as well as cells with a percentage of mitochondrial reads higher than 10%. The filtered data was then log-normalized and scaled using the Seurat package [v. 3.1.4, (Stuart et al., 2019)].

#### Definition of the T lymphocyte space

After filtering, gene expression data from PBMCs and enriched CD38^+^ T cells were log-normalized, merged, and scaled. We then performed PCA based on a subset of genes that exhibited high cell-to-cell variation (FindVariableFeatures Seurat function), constructed a KNN graph based on the Euclidean distance in PCA space considering the first 15 PCs (FindNeighbors Seurat function), and clustered the cells with the Louvain algorithm implemented in FindClusters Seurat function, with a resolution of 0.5.

Clusters from merged CD38^+^ T cell and PBMC libraries that contained mainly cells from PBMC libraries were excluded from the T cell space. T cell clusters were then confirmed by their high expression of *CD3D*, *CD3E*, and *CD3G* genes. At least 80% of the cells in the selected T cell clusters had a V(D)J sequence, while excluded clusters had a maximum of 10% of cells with a V(D)J sequence.

#### T cell clusters

After selecting T cells from PBMCs and enriched CD38^+^ T cells, we performed PCA based on a subset of genes that exhibited high cell-to-cell variation (FindVariableFeatures Seurat function). We then constructed a KNN graph based on the Euclidean distance in PCA space considering the first 15 PCs (FindNeighbors Seurat function), and clustered the cells with the Louvain algorithm implemented in FindClusters Seurat function, with a resolution of 0.5. UMAP was computed with the first 15 PCs.

#### Cluster annotation and statistical test

The 16 T cell clusters were manually annotated according to a set of T cell hallmark genes (depicted in Figure 2B). The specific name was assigned according to the Z-score standardized gene expression level of a particular set of genes (e.g. *MKI67*^++^/*FCGR3A*^++^) or well-established cellular states (e.g. CD4 naive). The complete processing, dimensionality reduction and clustering was performed with the Seurat package [v. 3.1.4, (Stuart et al., 2019)].

The frequency of each cluster was calculated as the percentage of cells in each cluster for each patient. Statistical testing for the difference in the frequency of each cluster was calculated with adjusted Dunn’s post-hoc test (Benjamini-Hochberg) for clusters with significant Kruskal-Wallis test (adjusted p-value (Benjamini-Hochberg) < 0.05, depicted as KW* in e.g.: Figure. 2E and Figure S2A).

#### Gene Ontology (GO) Enrichment Analysis

For the identification of differentially expressed genes between disease groups, we used pseudobulk gene expression, i.e., the sum of the raw counts from all cells in each patient among selected clusters of interest. The pseudobulk samples were then normalized according to the DESeq2 pipeline ((Love et al., 2014), v. 1.30.0). For further enrichment analysis, we selected differentially expressed genes with high counts (“baseMean” > 100) and p-value lower than 0.05. Differentially expressed genes were identified with the R package “DESeq2” v. 1.30.1, and GO enrichment analysis was performed with the R package “enrichR” v. 3.0.

### Gene Set Enrichment Analysis (GSEA)

The log2-fold change of differentially expressed genes (with high counts, i.e., “baseMean” > 100) from DESeq2 was used to define the ranked gene list used for GSEA. We tested three signatures (listed in Table S2): (1) RESPONSE TO TYPE I INTERFERON (GO:0034340), DEFENSE RESPONSE TO VIRUS (GO:0051607), and cytotoxicity. Gene lists (1) and (2) were obtained from the Molecular Signatures Database (MSigDB) (Subramanian et al., 2005), and for the cytotoxicity signature we used 17 cytotoxicity - associated genes taken from T_CELL_MEDIATED_CYTOTOXICITY (GO:0001913) and (Zhang et al., 2020). GSEA was performed with the R package “fgsea” v. 1.16.0 with 1000 permutations for statistical testing.

### ScRNA-seq data analysis of Rhapsody data (cohort 2)

#### Definition of the T lymphocytes space

From the complete landscape of immune cells in the PBMCs dataset, according to the labels of the original manuscript (Schulte-Schrepping et al., 2020), CD4^+^ T cells, CD8^+^ T cells, Activated T cells, Prol. cells and NK cells were selected for downstream analysis. The subset was scaled, centered and regressed against the number of detected transcripts per cell. PCA was calculated on the top 2000 most variable genes using the vst method integrated in Seurat’s FindVariableFeatures function and UMAP dimensionality reduction was performed. In the newly defined cellular space, we identified unwanted cells (non-T cells) according to the expression of lineage markers. With this approach, we removed NK cells and a small proportion of monocytes found to contaminate the original lymphocytes cluster. We further optimized the selection of the T cells space by removing cells that overlap with a recent study on NK cells, which utilizes the same dataset [Kramer at al. under revision]. The cleaned T cell space was once more scaled and PCA as well as UMAP dimensionality reduction were performed as described above. Cell clusters were calculated on the first 20 principal components (PCs) using Louvain clustering with a resolution of 1. All analysis steps were performed with Seurat [(Hao et al., 2020), v. 3.9.9.9032].

#### T cell cluster annotation

The 19 cell clusters were manually annotated according to a set of T cell hallmark genes (Figure 2B+S3B). Cell cluster names were assigned according to the expression levels of particular sets of genes (e.g. *MKI67^+^*^+^/*FCGR3A^+^*^+^) or well-established cellular states (e.g. CD4 naive)

#### Gene Set Enrichment Analysis (GSEA)

For the analysis of the differential regulation of specific pathways in moderate or severe COVID-19, GSEA was performed on pseudobulk samples. In this analysis, we included only samples from week 1 and 2 post-symptom onset. A pseudobulk sample, defined as the sum of the raw counts from all cells in each group, was generated for moderate and severe patients. The pseudobulk samples were then normalized according to the DEseq2 pipeline ((Love et al., 2014), v. 1.30.0). Log2-transformed fold change of each gene was calculated to define the ranked gene list used for GSEA. GSEA was performed with the fgsea package [v. 1.16.0] with 1000 permutations for statistical testing.

#### Data visualization

All the graphical visualization of the data was performed in R with the ggplot2 package [v. 3.3.3] with the exception of the heatmaps which were displayed using the pheatmap library [v. 1.0.12]

*Box plots:* Box plots are calculated in the style of Tukey, shortly the center of the box represent the median of the values, the hinges the 25th and 75th percentile and the whiskers are extended not further than the 1.5 * IQR (inter quartile range).

*Heatmap:* The heatmap shows the mean value of the scaled expression of each gene in each cluster.

*Dot plot:* The dot plot of the signature genes shown if figure S3F was calculated according to the Dotplot Seurat function scaling the expression values by gene.

#### Statistical testing

To calculate the statistical significance of the changes observed in the frequency of selected T cell clusters we first performed a Kruskal-Wallis rank sum test with Benjamini-Hochberg correction for multiple testing when more than one test was performed. For the clusters showing statistically significant results, we performed a pairwise Dunn post-hoc test with Benjamini-Hochberg correction for multiple testing. An adjusted p-value < 0.05 was considered significant and annotated above the box plot as follow: < 0.05 *, < 0.01 **, < 0.001 ***.

### Detection of SARS-CoV-2-specific IgG and IgA antibodies

Detection of IgG and IgA to the S1 domain of the SARS-CoV-2 spike (S) protein were assessed by anti-SARS-CoV-2 S1 IgG ELISAs (Euroimmun AG, Germany), as described elsewhere (Schlickeiser et al., 2020). Serum samples were tested at a 1:101 dilution and optical density (OD) ratios were calculated by dividing the OD at 450 nm by the OD of a calibrator sample tested within each run. Therefore, the calculated OD ratios can be used as a relative measure for the concentration of IgA and IgG antibodies in the tested sample. For IgG and IgA, OD ratios of above 1.1 were considered to be reactive.

### *Ex vivo* functional analyses of T cells

#### Degranulation assay and C3a binding assay

Frozen PBMC were thawed using Benzonase-containing wash buffer (RPMI, 2% FCS, Pierce Universal Nuclease, 250U/mL) seeded at 0.25x10^6^/well and rested overnight in a humidified incubator. Subsequent to overnight rest, PBMC were washed and resuspended in complete medium (RPMI1640 (Gibco, Thermo Fisher Scientific, USA), 2mM(1% V/V) GlutaMAX (Gibco, Thermo Fisher Scientific, USA), 10mM (1% V/V) HEPES (Sigma-Aldrich, USA), 10mM (1%v/v) Sodium Pyruvate (Gibco, Thermo Fisher Scientific, USA), 10% v/v Fetal Calf Serum (Sigma-Aldrich, USA), 1%(V/V) Penicillin/Streptomycin (Bio&Cell, Germany), 1%(V/V) MEM Non-essential amino Acid solution (Sigma-Aldrich, USA). Cells were stimulated with MACSiBeads (Miltenyi Biotec, Germany) coated with Biotin conjugated CD16 antibody (clone 3G8, Biolegend, USA) or corresponding isotype control (clone MOPC21, Biolegend, USA) at a ratio of 1:10 (cell to particle). Beads were loaded with indicated antibodies according to protocol provided by the manufacturer. PBMC were cultured in a humidified incubator (37°C, 5% C0_2_) for 6h in the presence of 1x Brefeldin A (eBioscienceTM, Thermo Fisher Scientific, USA) and 1x Monensin (Biolegend, USA). LAMP1 directed antibody (CD107a-APC, clone H4A3, final dilution 1:40) was added to the culture for the whole incubation period. Subsequent to 6h incubation cells were subjected to surface and intracellular staining. Fc receptor-mediated unspecific binding of antibodies was blocked by preincubation with human TruStain FcX Fc receptor blocking solution (Biolegend, USA) for 10 min at 4°C. Subsequently, cells were stained for CD3 BV711 (clone UCHT1, Biolegend), CD4 BV421 (clone OKT4, Biolegend), CD8 FITC (clone RPA-T8, Biolegend), CD16 BV605 (clone 3G8, Biolegend), CD38-PEcy7 (clone HB7, Biolegend), HLA-DR BV785 (clone L243, Biolegend) for 30 min at 4 C°. The fixable viability dye (ZombieRed, Biolegend) was incorporated in the surface staining mix. Surface antibody staining is performed in DPBS (Gibco, Thermo Fisher Scientific, USA), 2mM EDTA (Sigma-Aldrich, USA, 0,5%BSA Miltenyi Biotec, Germany). Cells were fixed for 20 min at 4°C with BD Cytofix/Cytoperm solution (BD Biosciences, USA) and intracellularly stained for 30 min at 4°C for Granzyme B APC/Fire 750 (clone QA16A02, Biolegend, USA), Perforin PE (clone dG9, Biolegend), Granzyme K PerCP/Cyanine5.5 (clone GM26E7, Biolegend) or TNFα PerCP/Cyanine5.5 (clone Mab11, Biolegend). Intracellular staining was performed in 1x Perm/ Wash permeabilization buffer (BD Biosciences, USA).

Flow cytometric, live (DAPI^-^), non-naive T cells (CD3^+^, CD45RA^+/-^, CCR7^-^ CD4^+^ or CD8^+^), were rested overnight in complete medium and washed in cold staining buffer (DPBS 2mM EDTA, 0,5% BSA). Prior to surface marker staining, non-specific, Fc receptor-mediated staining was blocked by 10 min pre-incubation at 4°C with human TruStainFcX (Biolegend, USA) blocking solution or Fc receptor blocking solution (Miltenyi Biotec, Germany). Surface marker staining was performed for 30 min at 4°C. Binding of complement split product hC3a was tested by incubation of cells for 60 min at 4°C with 50nM synthetic human C3a labelled with AF647 (Almac, UK) subsequent to fixation and permeabilization with BD Cytofix/Cytoperm^TM^ (BD Biosciences, USA) solution. Staining with C3a-AF647 was performed in 1x Perm/Wash permeabilization buffer (BD Biosciences, USA).

#### Endothelial-T cell-co-cultures

Human pulmonary microvascular endothelial cells (HPMECs, Promocell, Germany, passage 4-8) were plated on 96-well plates (96W10idf PET, Applied Biophysics Inc., USA) and grown to confluency for 48-72 h. Cell impedance was quantified by electric cell impedance sensing (ECIS® Z-Theta Applied Biophysics Inc., USA) at 4000Hz every 60 sec. For co-cultivation experiments, media was replaced by Opti-MEM^TM^ (Gibco, Thermo Fisher Scientific, USA). After 1 h stabilization phase, Convanacalin A (10 µM) from Canavalia ensiformis (Jack bean, Sigma-Aldrich Chemie GmbH, Munich, Germany, anti-CD16 beads (5 beads / T cell), and 20,000 flow-sorted, non-naive CD8^+^ T cells were added and cell barrier integrity was monitored continuously for 24 h. Resistance was normalized for each individual well to the baseline before treatment.

#### T cell differentiation cultures and C3a neutralisation

Frozen PBMC were thawed, washed twice with RPMI 1640 medium containing 2 % FBS and 0,02 % nuclease and afterwards resuspended in MACS buffer (DPBS with 0.5 % BSA, 2 mM EDTA) followed by enrichment of CD3^+^ cells using the human Pan T cell enrichment kit, (Miltenyi Biotec, Germany) according to the manufacturer’s protocol. CD3^+^ cells were activated in round-bottom wells (20000 cells/96 well) with 5 µg/ml plate-bound anti-human CD3, soluble 1 µg/ml anti-human CD28 and 20 IU/ml IL-2 (Proleukin) and the following additives: 1) 20 % non-inactivated serum (AB serum or serum from mild or severe COVID-19 patients), 2) 20 % heat-inactivated AB serum (30 min, 56 °C) +/-human recombinant C3a (20 nM), 3) 20 % non-inactivated serum from mild or severe COVID-19 patients +/- anti-human C3a (20 µg/ml). After 7 days of culture, cells were harvested and stained for flow cytometry using Zombie UV™ fixable viability kit (Biolegend, USA) for live/dead discrimination, Beriglobin® (CSL Behring, USA) for blocking of unspecific antibody binding (3,2 mg/ml) and CD3 BV421 (clone UCHT1), HLA-DR BV785 (clone L243), CD8 FITC (clone RPA-T8), CD38 PE-Cy7 (clone HB7) and CD16 AF700 (clone 3G8) for surface staining (all Biolegend, USA). Foxp3 staining buffer kit (Miltenyi Biotec, Germany) was used according to the manufacturer’s directions for intracellular staining of Ki67 eFluor506 (clone SolA15, Invitrogen, USA). Cells were acquired at a CytoFLEX LX flow cytometer and data analysis was done using Kaluza Analysis Software (both Beckman Coulter, USA).

T cells were differentiated as described above and rested overnight in a humidified incubator 37°C, 5% C0_2_. 20,000 T cells were transferred to a 96 well round well plate, washed 1x and resuspended in complete medium. Blaer1 GFP-/- cells were added to a ratio of 1: 5 (T cell: Blaer1 cell) subsequent to labelling with humanized monoclonal CD38-directed antibody (Daratumumab). To this end, Blaer1 cells were labelled for 30 min at 4°C with 50µg/mL Daratumumab. T cells and Blaer1 cells were incubated in a humidified incubator (37°C, 5% C0_2_) for 6h in the presence of 1x Brefeldin A (eBioscienceTM, Thermo Fisher Scientific, USA) and 1x Monensin (Biolegend, USA). LAMP1-directed antibody (CD107a-APC, clone H4A3, final dilution 1:40) was added to the culture for the whole incubation period. Prior to 6h incubation, cells were centrifuged for 1 min at 350 rcf. Subsequently to 6h incubation, cells were subjected to surface and intracellular staining. Fc receptor-mediated unspecific binding of antibodies was blocked by preincubation with human TruStain FcX Fc receptor blocking solution (Biolegend, USA) for 10 min at 4°C. TrueStain FCX is furthermore added to the respective staining cocktails. Subsequently, cells were stained for CD3 PE (clone UCHT1, Biolegend), CD4 APC/Fire 750 (clone RPT-A4, Biolegend), CD8 BV605 (clone RPA-T8, Biolegend) and CD16 FITC (clone 3G8, Biolegend). The fixable viability dye (ZombieRed, Biolegend) was incorporated in the surface staining mix. Surface epitope staining was performed in DPBS (Gibco, Thermo Fisher Scientific, USA), 2mM EDTA (Sigma-Aldrich, USA, 0,5%BSA (Miltenyi Biotec, Germany). Cells were fixed for 20 min at 4°C with BD Cytofix/Cytoperm solution (BD Biosciences, USA) and intracellularly stained for 30 min at 4°C Perforin PEcy7 (clone dG9, Biolegend, USA), IFNγ BV785 (clone 4S.B3, Biolegend, USA) for 30 min at 4 C°. Intracellular staining was performed in 1x Perm/Wash permeabilization buffer (BD Biosciences, USA).

#### Quantification of cytokines, chemokines & complement split products

Interleukin-6 (IL-6), interleukin-8 (IL-8=CXCL8), tumor necrosis factor (TNF) and monocyte chemoattractant protein-1 (MCP-1=CCL2) levels of cell culture supernatants were determined using a Milliplex MAP human cytokine/chemokine magnetic bead panel kit (Merck Millipore, USA, HCYTOMAG-60k). The assay was performed overnight with half of the volume according to the manufacturer’s instructions. The median fluorescent intensities were quantified on a Bio-Plex 200 system (Bio-Rad Laboratories, Inc., USA) and the results were calculated using Bio-Plex-Manager 6.1 software (Bio-Rad Laboratories, Inc., USA).

Concentrations of the C3a and C5a activation products were measured in EDTA plasma from patients using commercially available enzyme-linked immunosorbent assays (ELISA) kits (HK354 (C3a) at a 1:1000 dilution, HK349 (C5a) at a 1:4 dilution, Hycult Biotech, the Netherlands) according to the manufacturer’s protocols.

#### Plasma / serum proteomics

Proteomics was conducted using a recently developed platform technology for plasma and serum proteomics. (Demichev et al., 2020a; Messner et al., 2020). In brief, plasma and serum samples were prepared for mass spectrometry through denaturan, reduction, alkylation and tryptic digestion in 96 well plates using liquid handling robotics, and cleaned using solid phase microextraction. The samples were separated using high-flow rate reversed phase liquid chromatography with an 1290 Infinity II LC System (Agilent Technologies, USA), and data acquired using a Triple TOF 6600 Hybrid mass spectrometer (Sciex Ltd, USA) running SWATH MS (Gillet et al., 2012). Data was processed using DIA-NN (Demichev et al., 2020b), and post-normalized to control batch effects using information obtained from the repeated measurement of quality control samples.

## KEY RESOURCES TABLE

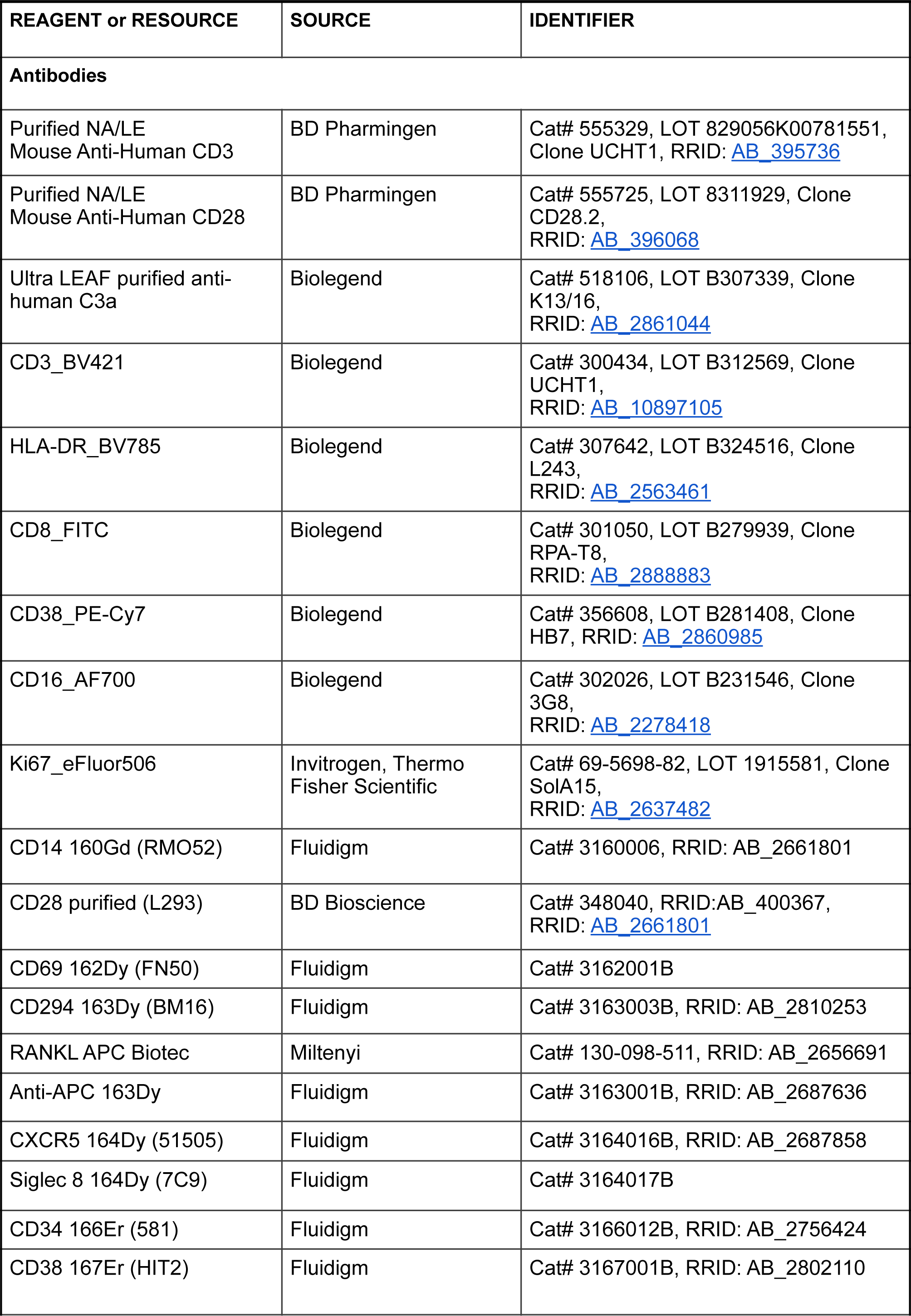
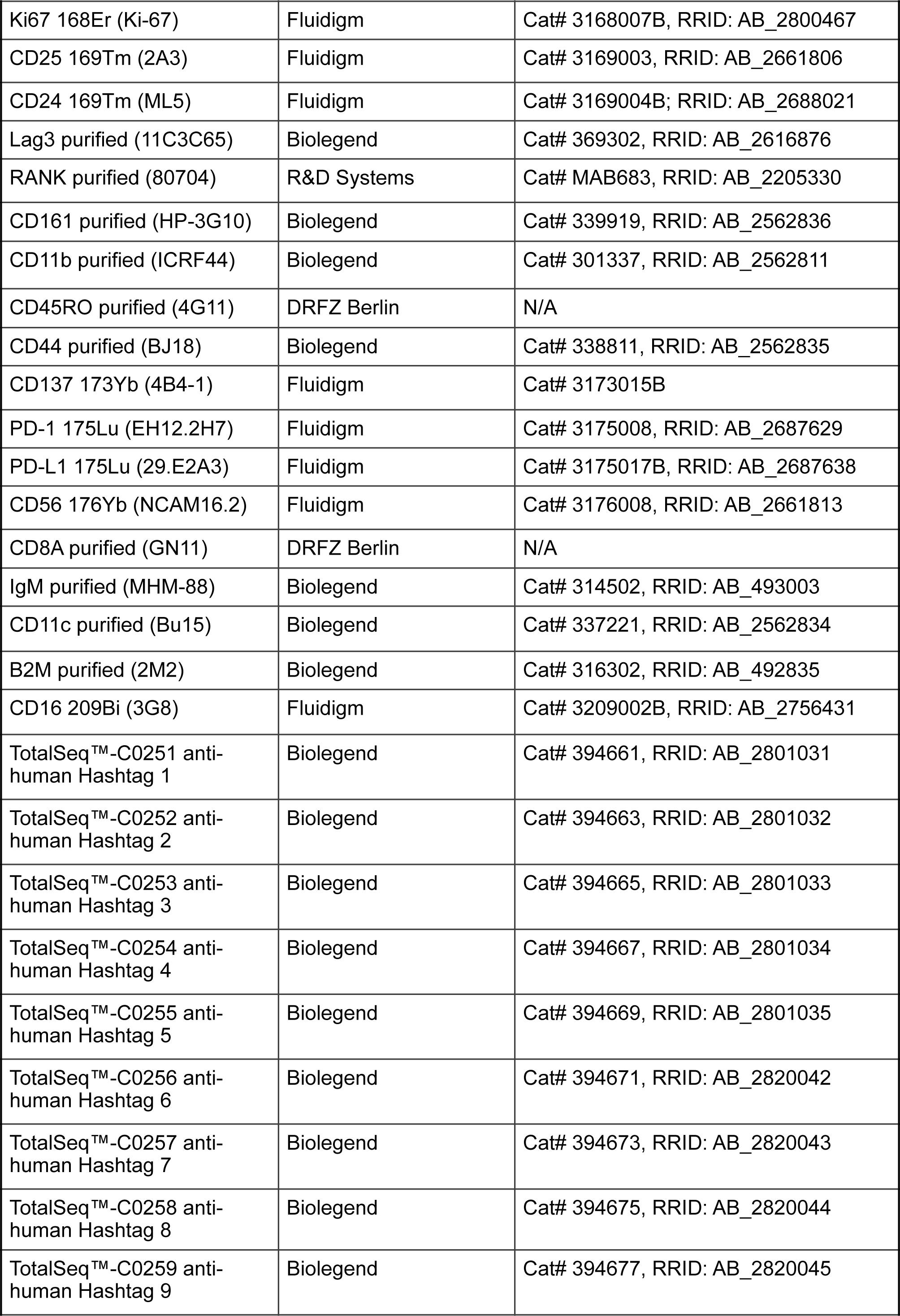
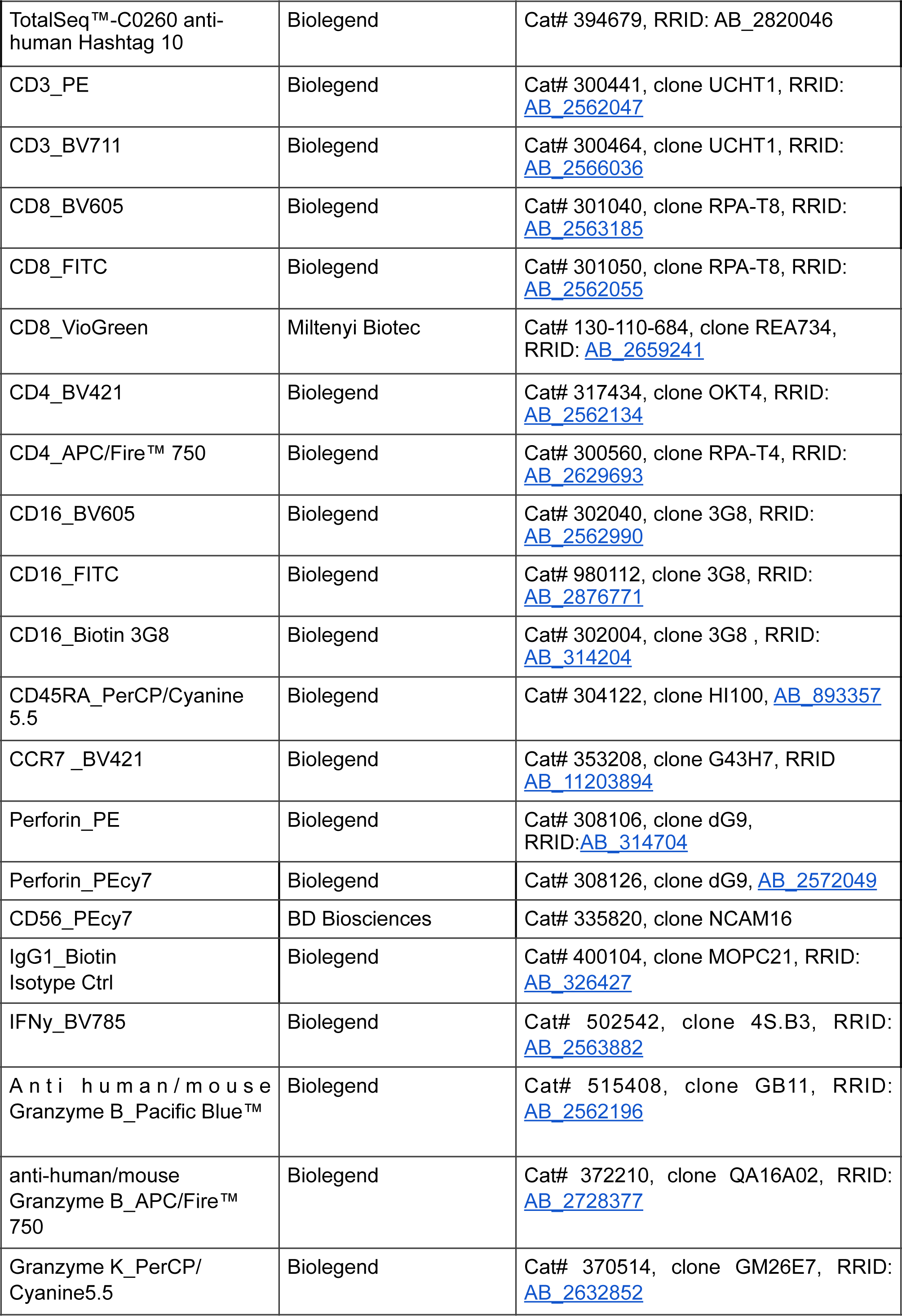
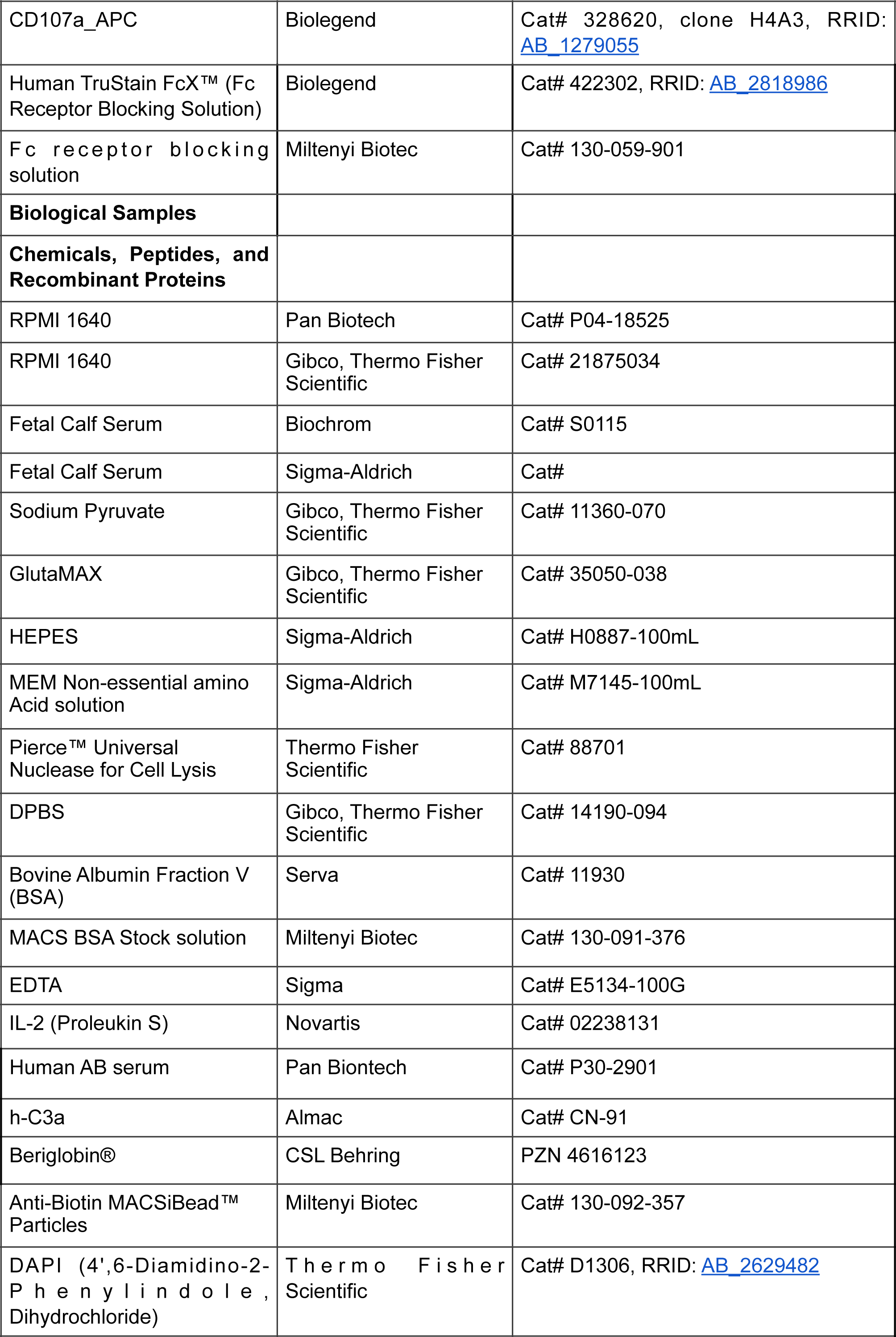
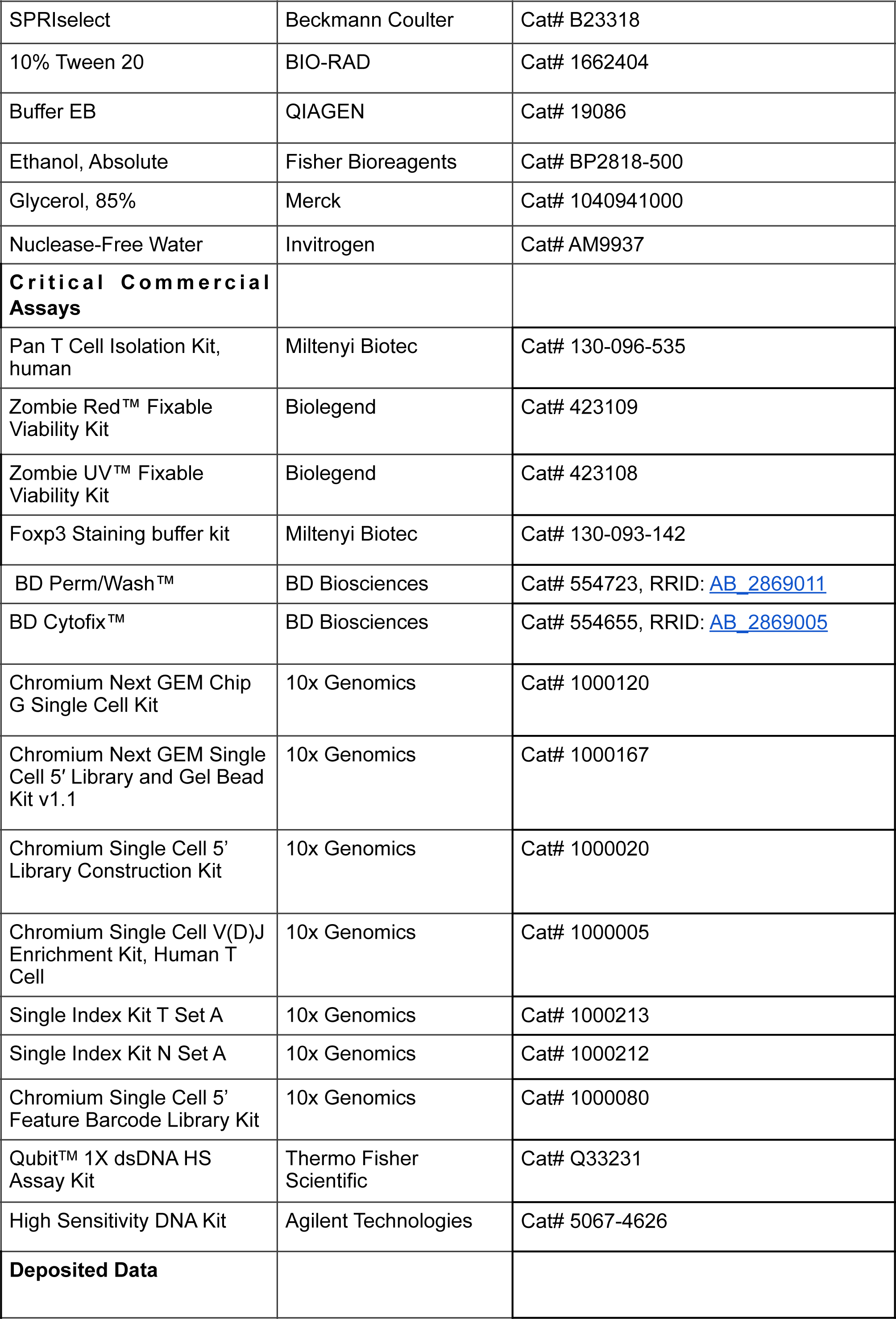
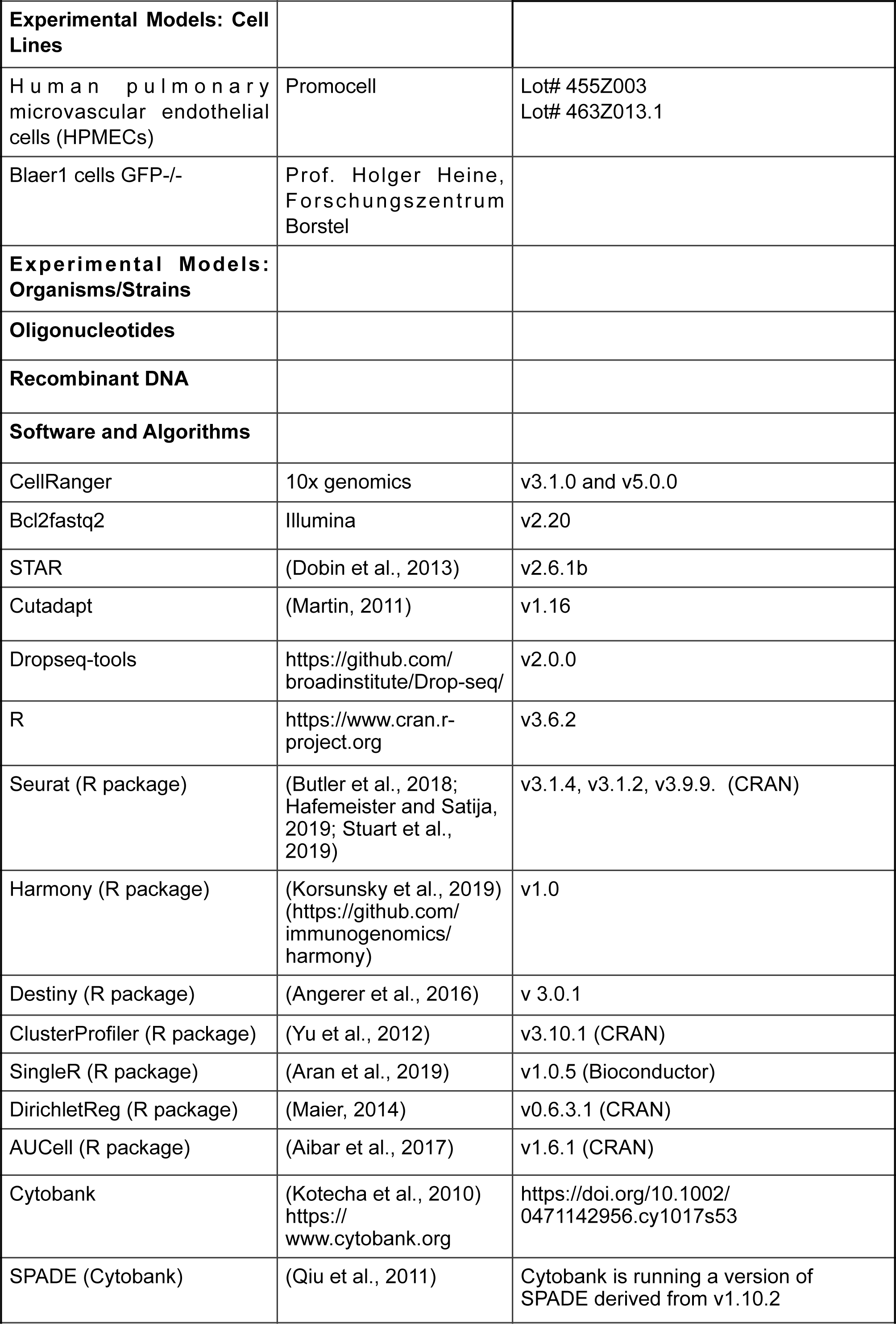
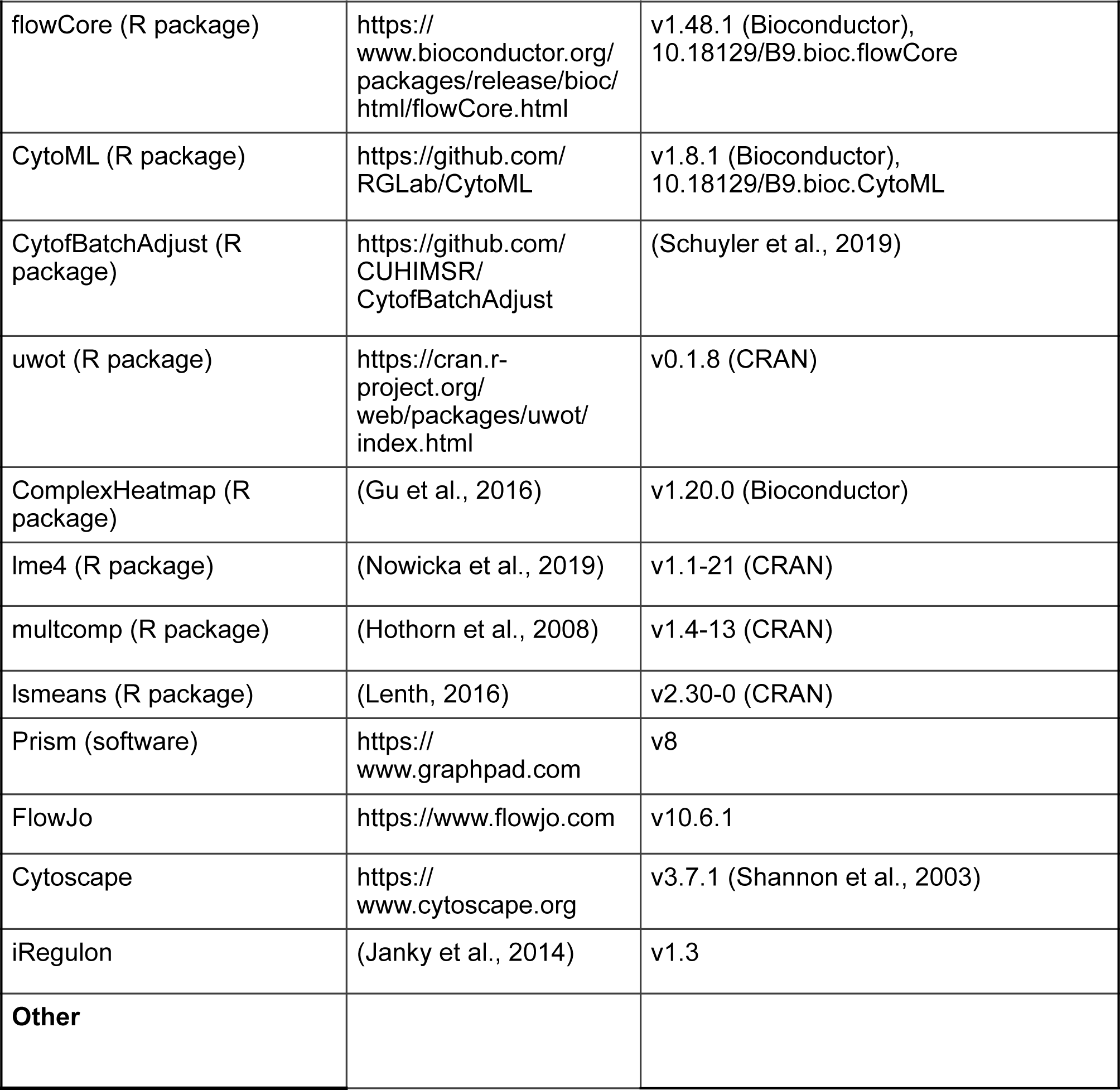

## Acknowledgements

We thank Katrin Vogt, Christine Appelt, Claudia Conrad, Anja Freiwald, Daniela Ludwig, and Vadim Fardzinov for technical support and Jonas Schulte-Schrepping, Elena de Dominico, Nico Reusch, Kristian Händler, Heidi Theis, Michael Kraut and Kevin Baßler for generating the scRNA-seq dataset of cohort 2. In addition, we thank Franziska Scheibe and Il-Kang Na for providing human antibodies.

We also thank Desireé Kunkel and Jacqueline Keye from the BIHFlow and Mass Cytometry Core facility for cell sorting and the BIH / MDC Genomics Platform for sequencing.

We thank all members of the Pa-COVID-19 collaborative study group:

Sascha S. Haenel, Mirja Mittermaier, Fridolin Steinbeis, Tilman Lingscheid, Bettina Temmesfeld-Wollbrück, Thomas Zoller, Daniel Grund, Christoph Ruwwe-Glösenkamp, Miriam S. Stegemann, Katrin M. Heim, Ralf H. Hübner, Bastian Opitz, Kai-Uwe Eckardt, Martin Möckel, Ulrike Bachmann, Felix Balzer, Claudia Spies, Marc Kastrup, Steffen Weber-Carstens, Frank Tacke, Christina Pley, Claudia Fink, Sarah Berger, Chantip Dang-Heine, Michael Hummel, Georg Schwanitz, Constanze Lüttke, Yinan Wu, Uwe D. Behrens, Maria Rönnefarth, Sein Schmidt, Christian Drosten, Martin Witzenrath, Alexander Krannich and Christof von Kalle for set up and realization of the study platform;

Linda Jürgens, Malte Kleinschmidt, Sophy Denker, Moritz Pfeiffer, Belén Millet Pascual-Leone, Luisa Mrziglod, Felix Machleidt, Sebastian Albus, Felix Bremer, Jan-Moritz Doehn, Tim Andermann, Carmen Garcia, Philipp Knape, Philipp M. Krause, Liron Lechtenberg, Yaosi Li, Panagiotis Pergantis, Till Jacobi, Teresa Ritter, Berna Yedikat, Lennart Pfannkuch, Christian Zobel, Ute Kellermann, Susanne Fieberg, Laure Bosquillon de Jarcy, Anne Wetzel, Christoph Tabeling, Markus C. Brack, Moritz Müller-Plathe, Jan M. Kruse, Daniel Zickler, Andreas Edel, Silke Leonhardt, Timur Özkan, Carola Misgeld, David Steindl, Marcel Wittenberg, Claas J. Steffen, Jan A. Graaw, Katharina Tielling, Ludwig Wiegank, Philipp Engelmann, Gottfried Lürzer, Victor Wegener, Stefan Angermair, Julia Heeschen, Moritz Weigeldt, Eike Wolter, Christoph Töpper, Anna Nothnagel, Sara Lange, Ralf F. Trauzeddel, Sebastian Stricker, Britta Stier, Roland Körner, Nils B. Müller, and Philipp Enghard for obtaining informed consent and biosamples;

Paula Stubbemann, Nadine Olk, Willi M. Koch, Alexandra Horn, Katrin K. Stoyanova, Saskia Zvorc, Yvonne Ahlgrimm, Wiebke Herud, Lucie Kretzler, Lil A. Meyer-Arndt, Linna Li, and Isabelle Wirsching for collection of clinical data;

Denise Treue*, Dana Briesemeister* and Jenny Schlesinger*, (*Central Biobank Charité/BIH; ZeBanC), for biobanking of samples.

This work was supported by the German Research Foundation (DFG): SA1383/3-1 to B.S.; SFB-TR84 114933180 to L.E.S., S.B., P.G., S.H. and W.M.K. INST 37/1049-1, INST 216/981-1, INST 257/605-1, INST 269/768-1, INST 217/988-1, INST 217/577-1, and EXC2151-390873048 to J.L.S.; GRK 2168 – 272482170, ERA CVD (00160389 to J.L.S.; SFB 1454 – 432325352 to A.C.A. and J.L.S.; SFB TR57 and SPP1937 to J.N.; GRK2157 to A.-E.S.; and ME 3644/5-1 to H.E.M.; RTG2424 to N.B.; SFB-TRR219 322900939, BO3755/13-1 Project-ID 454024652 to P.B.; the Berlin University Alliance (BUA) (PreEP-Corona grant to L.E.S. and V.M.C.); the Berlin Institute of Health (BIH) (to L.E.S., V.M.C.,B.S. and W.M.K.); Helmholtz-Gemeinschaft Deutscher Forschungszentren, Germany (sparse2big to J.L.S.), EU projects SYSCID (733100 to J.L.S.); European Research Council Horizon 2020 (grant agreement No 101001791 to P.B.); the DZIF, Germany (TTU 04.816 and 04.817 to J.N.); the Hector Foundation (M89 to J.N.); the EU projects ONE STUDY (260687), BIO-DrIM (305147) and INsTRuCT (860003) to B.S.); German Registry of COVID-19 Autopsies through Federal Ministry of Health (ZMVI1-2520COR201 to P.B.); Federal Ministry of Education and Research (DEFEAT PANDEMICs, 01KX2021 and STOP-FSGS-01GM1901A to P.B.); the Berlin Senate to German Rheumatism Research Centre (DRFZ); the Berlin Brandenburg School for regenerative Therapies (BSRT) to C.B.; the German Federal Ministry of Education and Research (BMBF) projects RECAST (01KI20337) to B.S., V.M.C., L.E.S and M.R.; VARIPath (01KI2021) to V.M.C.; NUM COVIM (01KX2021) to L.E.S., V.M.C., F.K., J.L.S., J.N. and B.S.; RAPID to and S.H.,; SYMPATH to N.S. and W.M.K.; PROVID to S.H. and W.M.K.; ZissTrans (02NUK047E) to N.B; National Research Node ‘Mass spectrometry in Systems Medicine (MSCoresys) (031L0220A) to M.R. and N.B.; Diet–Body–Brain (DietBB) (01EA1809A) to J.L.S.; the UKRI/NIHR through the UK Coronavirus Immunology Consortium (UK-CIC), the Francis Crick Institute through the Cancer Research UK (FC001134), the UK Medical Research Council (FC001134), the Wellcome Trust (FC001134 and IA 200829/Z/16/Z) to M.R.; a Charité 3R project (to B.S., S.H., W.M.K.); and an intramural grant from the Department of Genomics & Immunoregulation at the LIMES Institute to A.C.A.

We are grateful to the patients and donors volunteering to participate in this study making this research possible in the first place and wish for a speedy and full recovery.

## Author contributions

Conceptualization: M.R., J.L.S., A.C.A, F.K., L.E.S., N.B., B.S. Methodology: P.G., L.B., L.M., T.K., M.St., V.M.C., P.B., W.M.K.; Software/Formal analysis: R.A.G., L.B., V.D., I.G., S.K.A, V.M.C., P.B., N.B.; Investigation: P.G., L.B., S.B., L.M., T.K., C.G., M.S., M. St., J.S., H-P.D., D.P., M.M., P.B.; Resources: L.J.L., P.T.L., E.T.H., H.E.M., A.R.S., S.H., P.B., M.D., H.We., N.S., A.U., H.M.R., J.N., C.M., C.T., F.K.; Data Curation: D.B., E.W., M.L., B.O.W.; Writing – Original Draft: P.G., R.A.G., L.B., S.B., L.M., T.K., M.S., M.B., V.M.C., B.S.; Writing – Review & Editing: P.G., R.A.G., L.B., M.R., W.M.K., C.M., J.L.S., A.C.A. C.T., F.K., L.E.S., N.B., B.S.

## Supplemental figures, titles, and legends

**Supplemental Figure 1.**
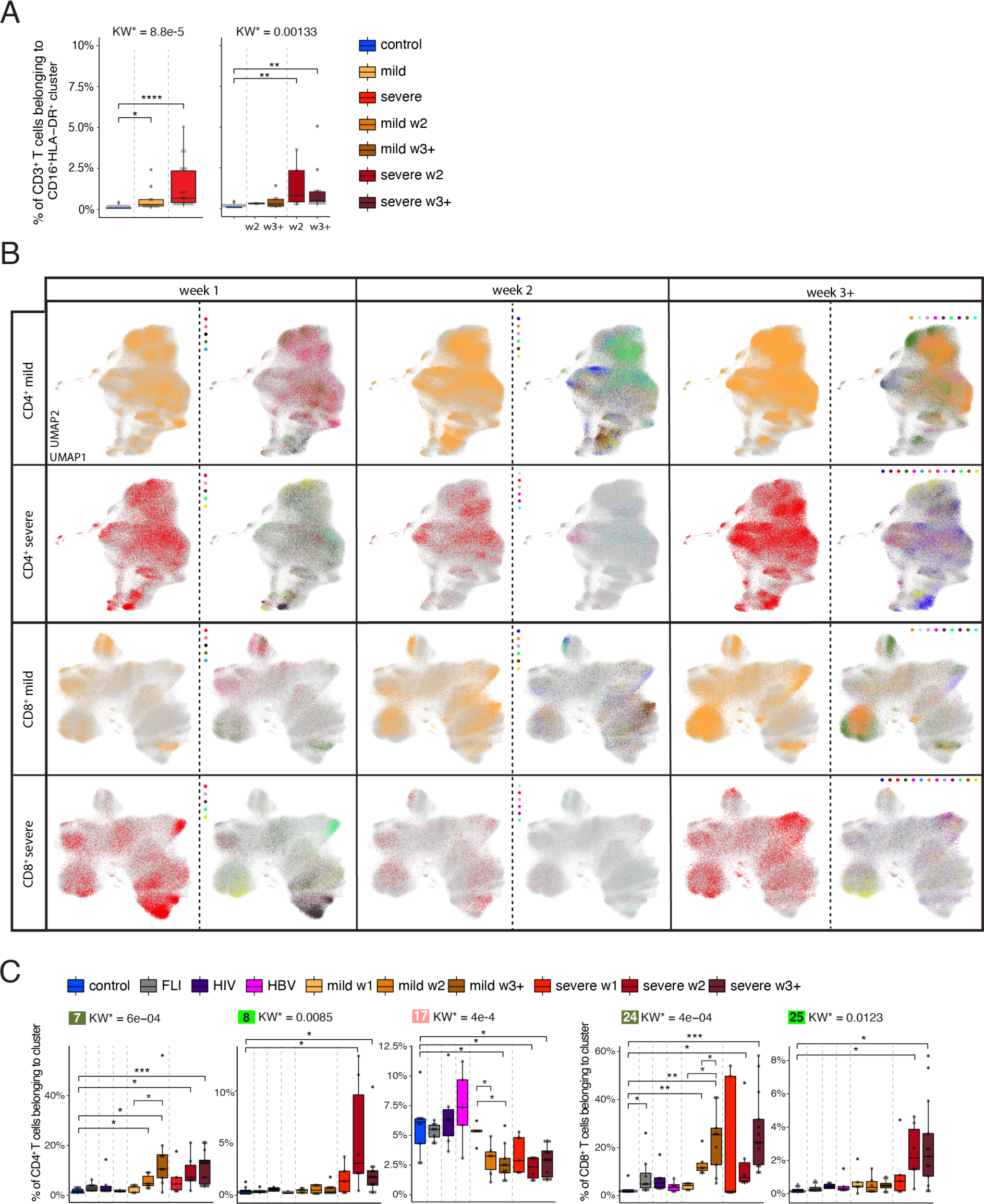
Weekly changes in CD4^+^ and CD8^+^ T cell cluster composition in mild versus severe COVID-19. **A**, Box plots of CD16^+^HLA-DR^+^ CD3^+^ T cells determined by flow cytometry (cohort 2) of samples from controls (n = 11) as well as mild COVID-19 acute (n = 11) and severe COVID-19 acute (n = 20) patients collected during the acute infection (all samples collected, left panel) or samples collected during week two and three post-symptom onset only (right panel). KW* shows the adjusted p-value (Benjamini-Hochberg) of a Kruskal-Wallis test. The abundance of each cluster was compared between severity groups via adjusted Dunn’s post-hoc test (Benjamini-Hochberg) for clusters with KW* < 0.05. All combinations where tested, only comparisons with healthy controls are shown. (*p<0.05, **p < 0.01, ***p < 0.001). **B,** UMAPs generated of CD4^+^ and CD8^+^ T cells from mass cytometry data of samples from COVID-19 patients collected during week one, two and three post-symptom onset. Cells are colored according to (left) disease severity (yellow, mild COVID-19 acute phase; red, severe COVID-19 acute phase), and (right) patient ID. **C,** Box plots of CD4^+^ (7, 8, 18) and CD8^+^ (24, 25, 32) T cell clusters determined by mass cytometry (whole blood, cohort 1) of samples from controls (n = 9), FLI (n = 8), HIV (n = 6), HBV (n = 5) as well as acute mild COVID-19 week 1, (n = 5), week 2 (n = 6), week 3+ (n = 9), and acute severe COVID-19 week 1 (n = 5), week 2 (n = 6), week 3+ (n = 13) patients collected during week one, two and three post-symptom onset. KW* shows the adjusted p-value (Benjamini-Hochberg) of a Kruskal-Wallis test. The abundance of each cluster was compared between severity groups via adjusted Dunn’s (Benjamini-Hochberg) for clusters with KW* < 0.05. All combinations where tested, only comparisons with healthy controls and within COVID-19 disease are shown. (*p<0.05, **p < 0.01, ***p < 0.001).

**Supplemental Figure 2.**
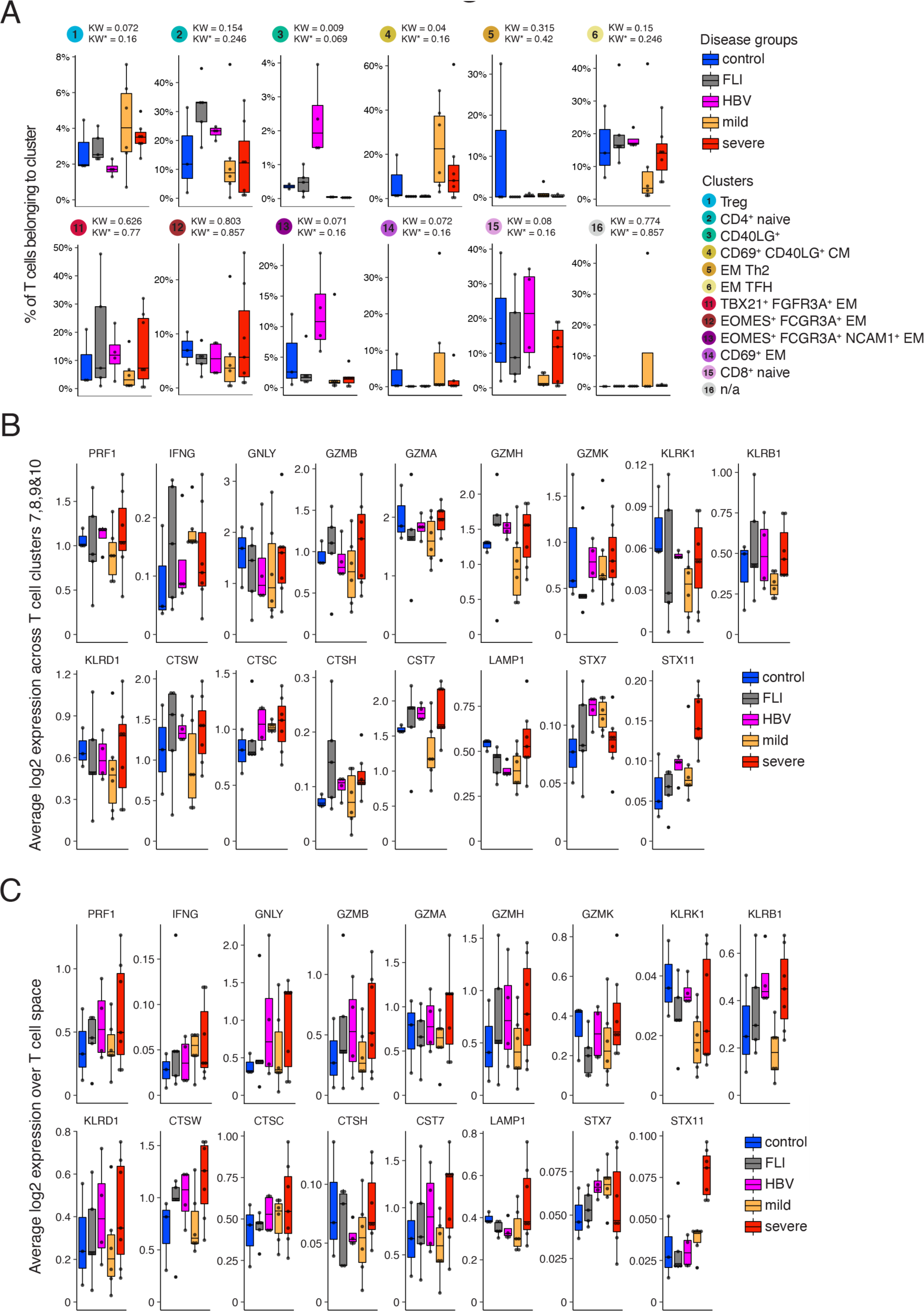
scRNA-seq T cell clusters and their cytotoxic gene signature in samples from COVID-19 patients or patients with other infections. **A**, Box plots of the percentage of cells in the remaining scRNA-seq T cell clusters generated from controls (n = 3), FLI (n = 5), HBV (n = 4), mild COVID-19 (n = 6) and severe COVID-19 (n = 7) patient samples of cohort 1. KW* shows the adjusted p-value (Benjamini-Hochberg) of a Kruskal-Wallis test. **B**, Box plots of the average log2-transformed expression of all genes defining the cytotoxicity gene signature in cells belonging to clusters 7,8,9 & 10, generated from controls (n = 3), FLI (n = 5), HBV (n = 4), mild COVID-19 acute phase (n = 6) and severe COVID-19 acute (n = 7) patient samples of cohort 1. **C**, Box plots of the average log2-transformed expression of all genes defining the cytotoxicity gene signature in cells belonging to all T cell clusters generated from controls (n = 3), FLI (n = 5), HBV (n = 4), mild COVID-19 (n = 6) and severe COVID-19 (n = 7) patient samples of cohort 1.

**Supplemental Figure 3.**
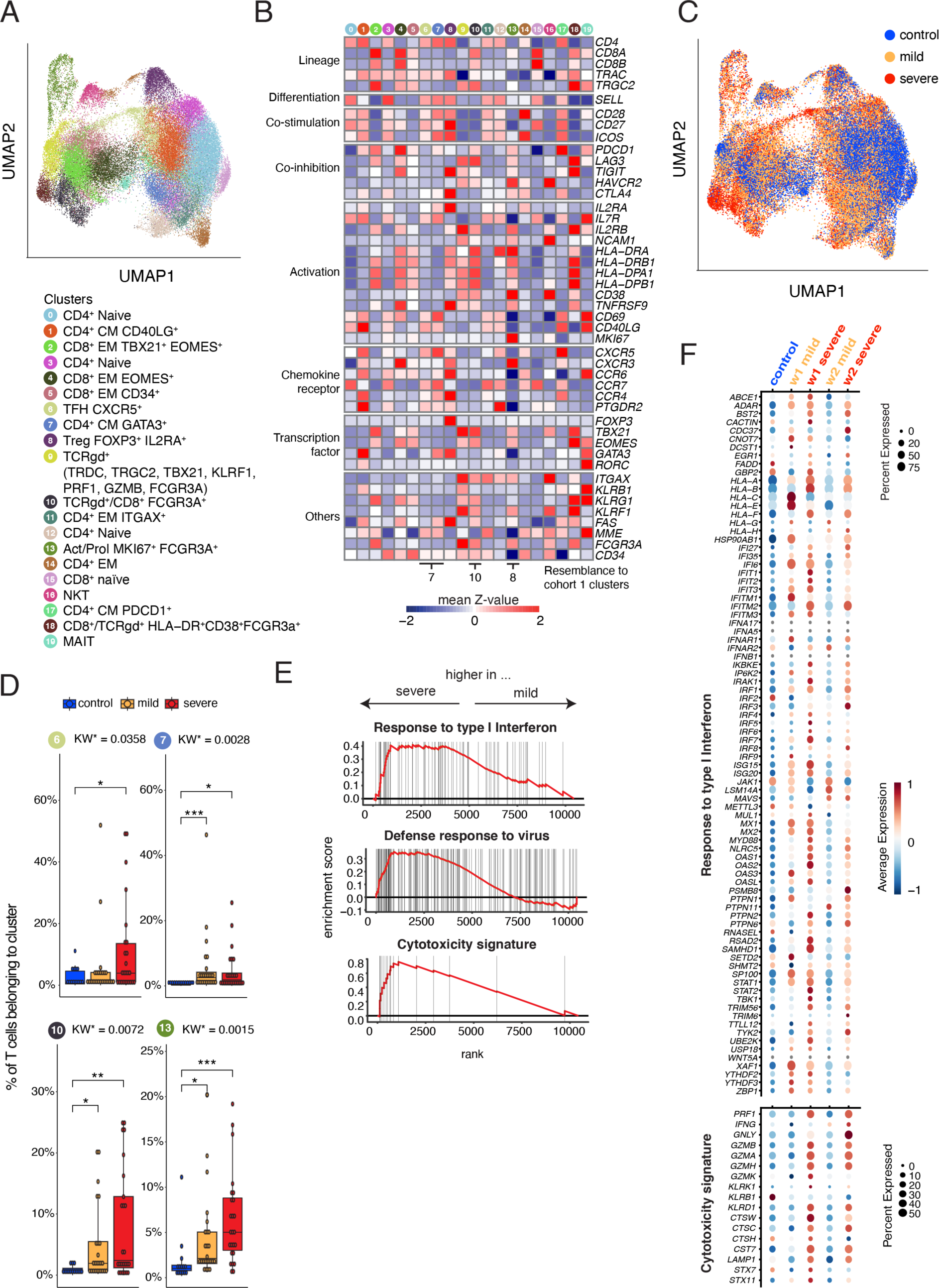
scRNA-seq T cell cluster and their cytotoxic gene signature in samples from COVID-19 patients or controls of cohort 2. **A**, UMAP of the T cell subset from the PBMC dataset of Schulte-Schrepping et al. (Schulte-Schrepping et al., 2020), including controls (n = 13), mild COVID-19 (n = 21) and severe COVID-19 (n = 29) patients. **B**, Heatmap of selected marker expression of the T cell subset from the PBMC dataset of Schulte-Schrepping et al. (Schulte-Schrepping et al., 2020), including controls (n = 13), mild COVID-19 (n = 21) and severe COVID-19 (n = 29) patients. **C,** UMAP of T cell clusters as shown in A with cells coloured according to disease group origin: blue, age-matched controls (n = 13); yellow, mild COVID-19 acute phase (n = 21); red, severe COVID-19 acute phase (n = 29). **D**, Box and whisker (10–90 percentile) plots of a selection of cohort 2 scRNA-seq T cell clusters, generated from controls (n = 13), mild COVID-19 acute phase (n = 21) and severe COVID-19 acute phase (n = 29) patient samples. Selected clusters show a TFH phenotype (cluster 6 & 7) or display *FCGR3A* expression and increased frequency in severe COVID-19 (cluster 10 & 13) generated from controls (n = 13), mild COVID-19 acute phase (n = 21) and severe COVID-19 acute phase (n = 29) patient samples. KW* shows the adjusted p-value (Benjamini-Hochberg) of a Kruskal-Wallis test. The abundance of each cluster was compared between severity groups via adjusted Dunn’s post-hoc test (Benjamini-Hochberg) for clusters with KW* < 0.05. All combinations where tested, only comparisons with healthy controls are shown. (*p<0.05, **p < 0.01, ***p < 0.001). **E**, GSEA performed on the ranked gene list of the comparison severe vs. mild COVID-19. The graph shows the mapping of the signature genes on the ranked gene list. The curve corresponds to the running sum of the weighted enrichment score (ES). The ranked gene list was calculated from the normalized pseudobulk expression data of severe and mild samples week 1 and 2 post symptoms onset across clusters 6, 7, 10 & 13. **F**, Dot plot of the expression of the genes included in the “Cytotoxicity” and “Response to type I interferon” signatures in control, mild COVID-19 and severe COVID-19 samples in week 1 and 2 post symptoms onset across clusters 6, 7, 10 & 13. The dots are colored by the scaled gene expression across the groups and the size is proportional to the ratio of cells expressing the specific gene.

**Supplemental Figure 4.**
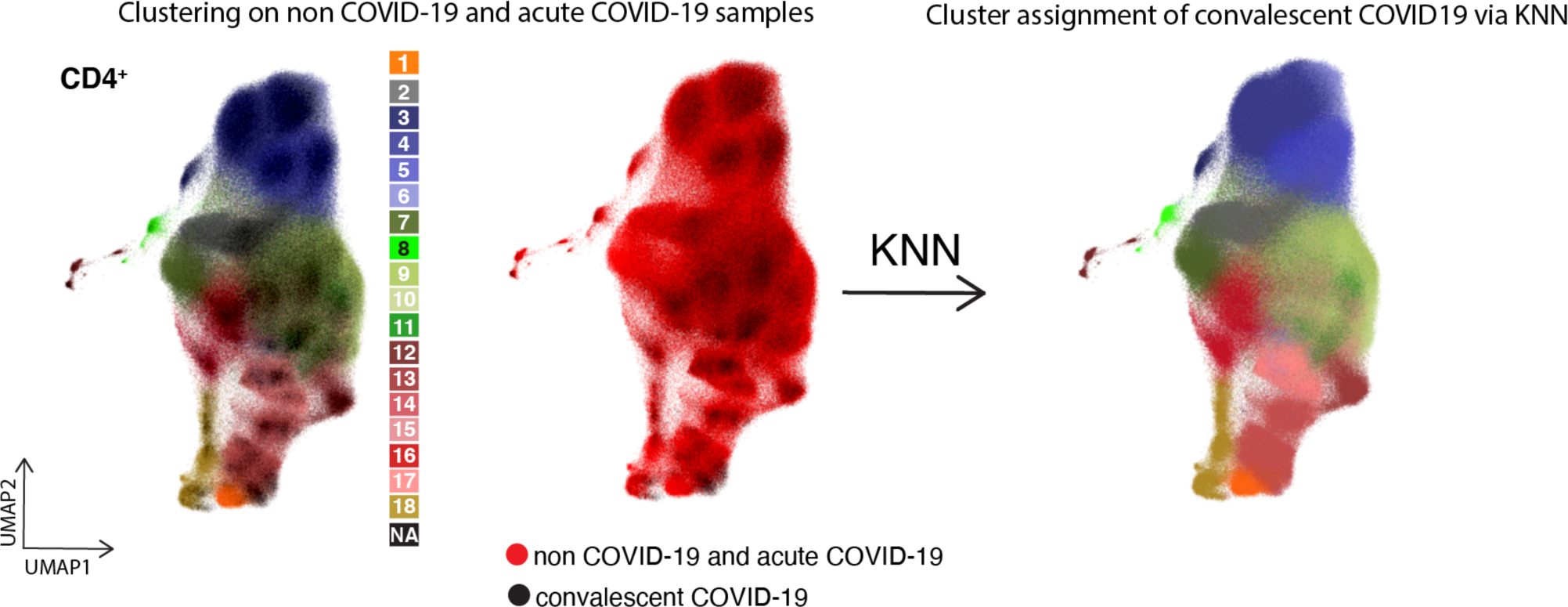
Exemplary graph visualizing the assignment of CD4^+^ T cells measured during the convalescent phase to CyTOF T cell clusters identified during acute COVID-19. **A**, UMAP generated with CD4^+^ T cells from mass cytometry data, coming from acute (non-CV19 and CV19) and convalescent CV19 samples. Left: Cells from acute samples are colored according to the cell cluster origin (see legend), while cells from convalescent CV19 samples haven’t been assigned to a specific cluster (NA). Middle: Cells from acute and convalescent CV19 samples are colored in red and black, respectively. Right: Cells from acute and convalescent CV19 samples are colored according to the cell cluster, after assigning clusters to cells from convalescent CV19 samples via KNN approach.

## Supplemental Tables

**Table S1.** Cohort details.

**Table S2.** Gene lists for the signatures “Response to Type I Interferon”, “Defense Response to virus” and “Cytotoxicity” used for GSEA.

